# Haplotype Analysis Reveals Pleiotropic Disease Associations in the HLA Region

**DOI:** 10.1101/2024.07.29.24311183

**Authors:** Courtney J. Smith, Satu Strausz, FinnGen, Jeffrey P. Spence, Hanna M. Ollila, Jonathan K. Pritchard

## Abstract

The human leukocyte antigen (HLA) region plays an important role in human health through involvement in immune cell recognition and maturation. While genetic variation in the HLA region is associated with many diseases, the pleiotropic patterns of these associations have not been systematically investigated. Here, we developed a haplotype approach to investigate disease associations phenome-wide for 412,181 Finnish individuals and 2,459 traits. Across the 1,035 diseases with a GWAS association, we found a 17-fold average per-SNP enrichment of hits in the HLA region. Altogether, we identified 7,649 HLA associations across 647 traits, including 1,750 associations uncovered by haplotype analysis. We find some haplotypes show trade-offs between diseases, while others consistently increase risk across traits, indicating a complex pleiotropic landscape involving a range of diseases. This study highlights the extensive impact of HLA variation on disease risk, and underscores the importance of classical and non-classical genes, as well as non-coding variation.

## Introduction

The major histocompatibility complex (MHC) plays a crucial role in mediating tissue graft compatibility and immune system recognition of pathogens and self [1–3]. The human MHC, referred to as the human leukocyte antigen (HLA) region, has been found to be associated with numerous diseases [2–8]. The tension between being able to recognize a diverse array of pathogens while avoiding autoimmunity suggests that variants within the HLA region may affect multiple distinct phenotypes simultaneously. Yet, little work has been done to characterize the patterns of pleiotropy and the trade-offs across diseases within the region.

The HLA region is approximately 5 megabases in length, and contains hundreds of genes, but is most known for the classical HLA genes, which are involved in response to infection and autoimmunity [9]. The classical HLA genes, which include class I genes (*HLA-A*, –*B*, –*C*) and class II genes (*HLA-DR*, –*DQ*, and –*DP*), encode cell surface proteins that present peptides to immune cells resulting in activation and maturation [10].

The classical HLA genes are highly polymorphic, with each gene having multiple distinct alleles. These alleles are functionally diverse: some act as generalists, and others are specific to particular types of peptides [11–13]. Different HLA alleles vary in their ability to recognize certain pathogens, thus genetic variation modulating this ability can result in a variety of disease associations [9, 14]. Meanwhile, some pathogens have evolved to avoid common HLA alleles in a host-pathogen arms race [15, 16]. This arms race has resulted in long-term balancing selection at classical HLA genes, leading to trans-species polymorphisms and extreme nucleotide diversity—more than 70-times the genome-wide average [17–19].

At the individual level, this genetic variation in the classical HLA genes affects the ability of the immune system to detect pathogens, fight infections, and attack cancerous cells, as well as the ability to limit inappropriate immune responses, such as autoimmune diseases [2–5]. Furthermore, genetic variation in the HLA region can influence the balance between these conflicting goals of pathogen response and the prevention of autoimmunity, resulting in potential risk trade-offs [20–22]. On the other hand, the risk trade-offs between autoimmunity, infection, and other traits can be more complicated, as demonstrated by Epstein-Barr virus (EBV) infection. Chronic EBV infection is known to cause various cancers, including nasopharyngeal carcinoma and Hodgkin lymphoma [23–25], and it has also been shown to play a role in the development of multiple sclerosis, a degenerative demyelinating disease of the central nervous system caused by immune-mediated inflammation [26, 27]. Although there is clinical evidence of the complex interplay between infection, autoimmunity, cancer, and other diseases, the genetic contribution to these disease trade-offs and risks has not been well-characterized at the biobank level [2, 20, 21, 23].

Association studies have implicated particular HLA alleles in many diseases [6, 7]. These canonical HLA association studies have provided countless biologically and clinically informative associations, for example, seronegative spondyloarthritis has been associated with the *HLA-B27* allele family, Type 1 Diabetes with the *HLA-DR3* allele family, and Rheumatoid arthritis with the *HLA-DR4* allele family [28, 29]. In addition to providing biological insight into disease mechanisms, these studies have resulted in the use of HLA allele associations in the clinical setting [30–32].

While there has been much focus on protein-coding variation within the classical HLA genes, there has been less work characterizing the majority of the genetic variation in the region, which falls outside of the coding regions of the classical HLA genes. Disease-associated variants are typically presumed to be protein-coding, affecting the peptide-binding groove of a classical HLA gene, but variation in regulatory regions may also be a major risk factor in a subset of diseases by influencing gene expression [33–35]. Recent experimental studies have demonstrated that for some traits, regulatory variation in the region confers more risk than HLA coding variation [36]. There is also evidence for disease associations with variation in non-HLA genes within the locus, including *C4A* [37], *SLC44A4* [38], and *NOTCH4* [39]. Therefore, investigation of genetic variation throughout the entire HLA region has the potential to reveal additional contributions beyond those found by HLA allele analysis alone.

Analyses of the HLA region in genome-wide association studies (GWAS) in large cohorts such as FinnGen [40], UK Biobank [41], and Japan Biobank [42] have identified many trait associations with single nucleotide polymorphisms (SNPs) in the HLA region [43]. These traits span a variety of systems, including infections such as HIV [44] and Hepatitis B [45], and autoimmune conditions ranging from neurological conditions (such as multiple sclerosis [46]), gastrointestinal disorders (such as Celiac disease and inflammatory bowel disease [47]), and rheumatic disorders (such as systemic lupus erythematosus [48]). These studies typically either investigate associations with many traits across the entire genome [8, 39, 49], treating the HLA region as just another locus, or they specifically focus on the HLA region but consider only a small number of traits at a time [50, 51]. However, in order to understand how genetic variation in the HLA region contributes to the complicated interplay between different disease risks, it is crucial to study associations for many traits simultaneously. This motivates the need for investigating the role of HLA loci in modulating trade-offs in these disease associations at the phenome-wide scale.

In this study, we quantified how genetic variation and pleiotropy at the HLA region contribute to disease risk across a broad range of diseases. We analyzed data from 412,181 Finnish individuals for 2,459 traits. We focused on understanding the spatial distribution of disease associations throughout the HLA region and the nature of pleiotropy between different traits. We developed a haplotype-based approach to robustly characterize patterns of disease associations throughout the entire HLA region, including non-coding variation and variation outside of classical HLA genes. We applied our approach at a phenome-wide scale and evaluated the role of HLA in modulating risk and trade-offs across a broad range of diseases in the context of the full complexity and breadth of HLA genetic variation.

## Results

### Enrichment of significant trait associations in the HLA region

To identify disease associations with genetic loci throughout the entire HLA region, we analyzed data from 412,181 Finnish individuals and 2,459 traits (Figure 1). We used fine-mapped GWAS summary statistics released by FinnGen, as well as new association data we generated at the level of individual phased haplotypes and HLA alleles. We corrected for sex, age, and the first ten principal components of the genome-wide genotype matrix (see Methods). Results from these association tests were used in subsequent analyses.

**Figure 1:**
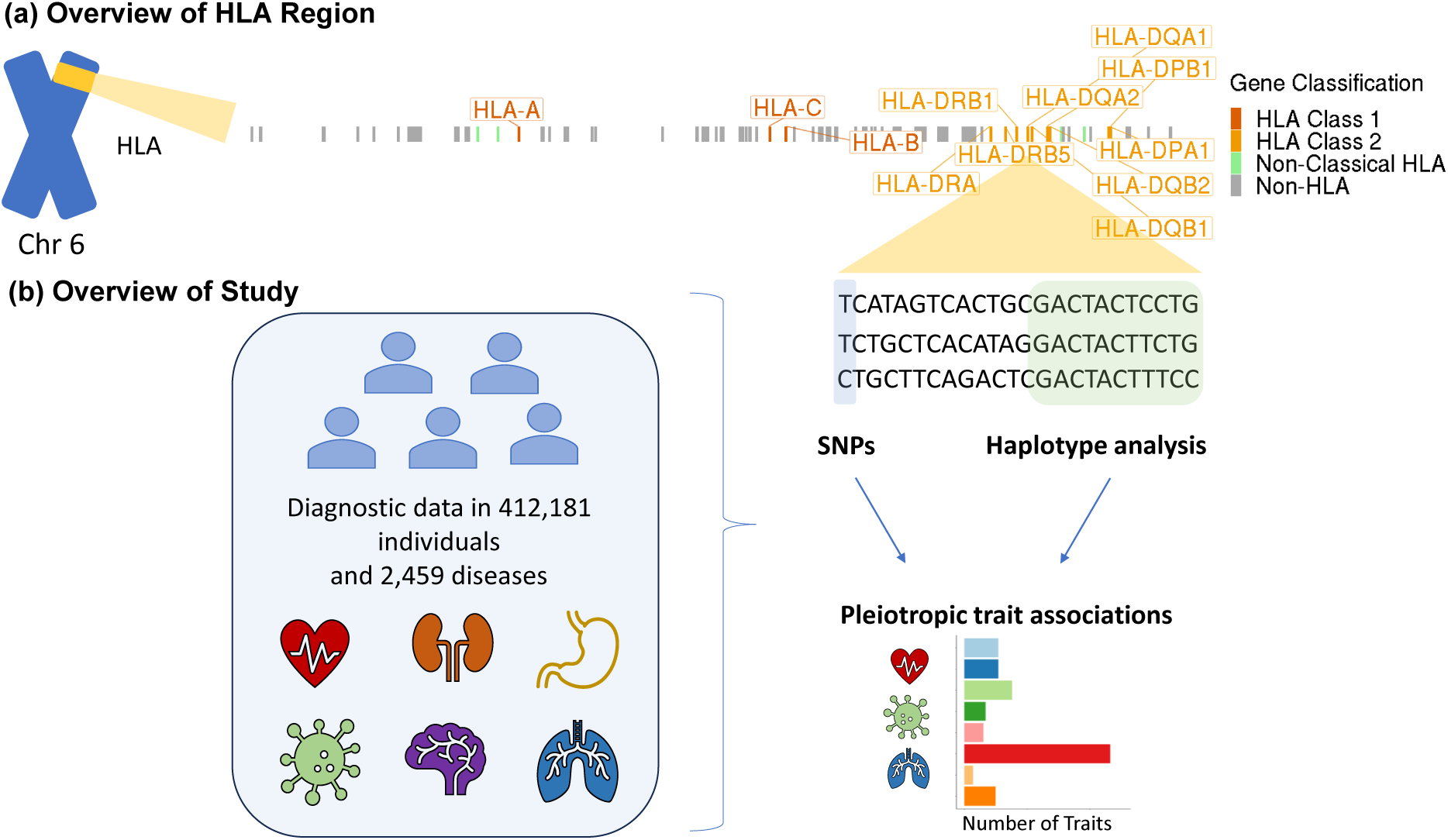
Study Overview. **A**. An overview of the HLA region showing the nearest genes to trait-associated SNPs, colored by HLA class, spanning approximately 5 megabases. **B**. An overview of the study data and design.

While the importance of HLA variation in disease has been well-established, we first sought to systematically quantify the enrichment of association signals across diseases, focusing on how enrichment varies by disease type. We considered the 1,035 disease traits in FinnGen that had at least one genome-wide significant association anywhere in the genome. We then identified independent genome-wide significant SNP associations for each trait, and binned these SNPs into 100 kb bins (Figure 2a). We found the mean number of significant associations per bin was 2.75, with a median of 1. One of the bins on chromosome 6 that overlaps the class II region of the HLA region had the highest number of associations in a single bin with 282 associations. Five of the six bins with the most associations overlapped the HLA region. The remaining bin is on chromosome 19 and has 101 associations. This bin contains an apolipoprotein gene cluster including *APOE*, *APOC1*, *APOC2*, *APOC4*, which are involved in lipid metabolism and affect Alzheimer’s disease risk. These results show that the HLA region harbors a higher density of disease associations than the rest of the genome.

**Figure 2:**
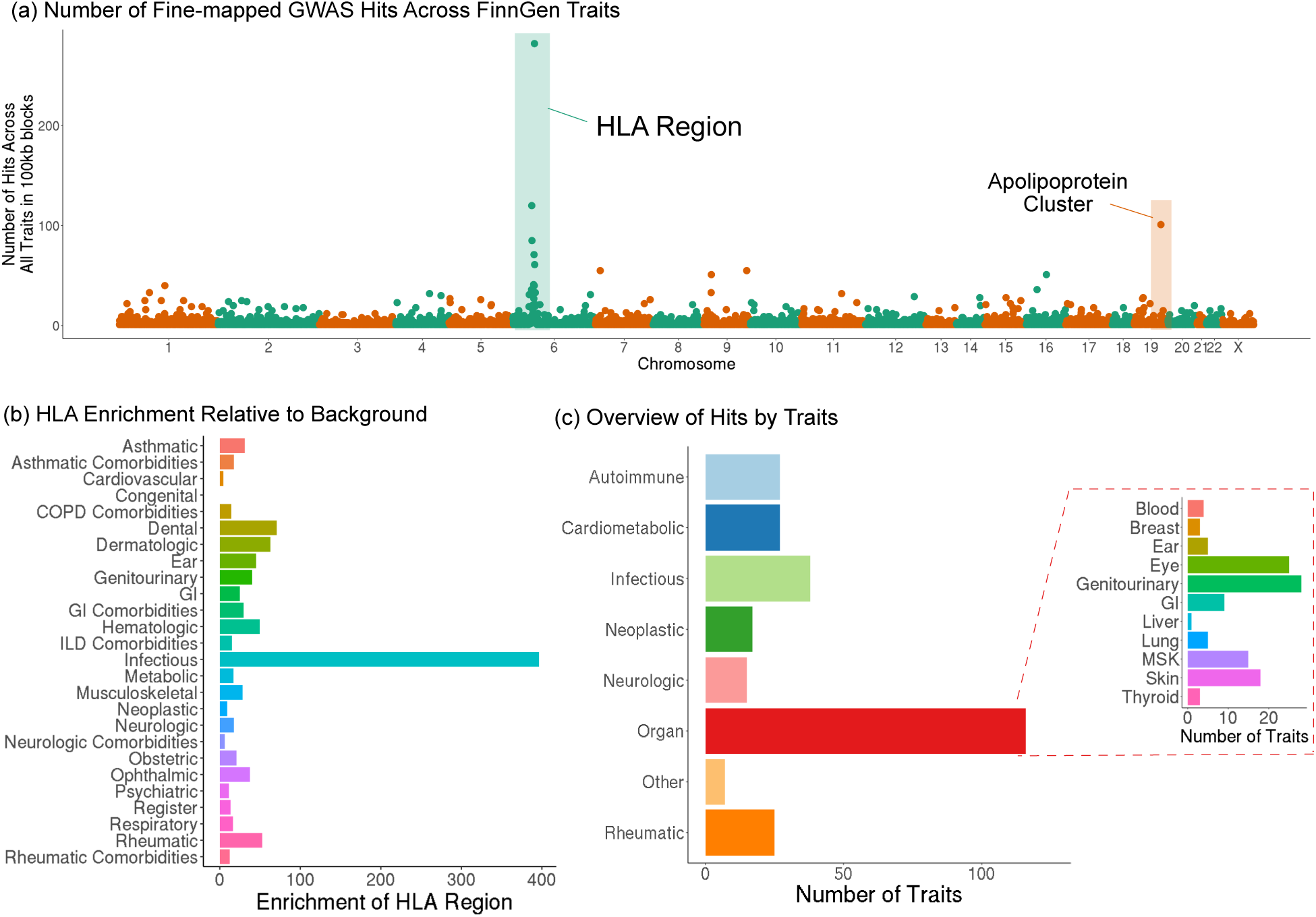
Distribution of GWAS hits across the genome and trait group enrichment. **A**. Distribution of fine-mapped GWAS hits throughout the genome across 1,035 FinnGen disease traits, binned into 100 kb bins. **B**. Enrichment of association signal in the HLA region by disease group. The 1,035 diseases were categorized into 45 disease groups based on ICD codes and the average per SNP enrichment in the HLA region was calculated by comparing the number of independent associations in the HLA region relative to that in the rest of the genome. **C**. Classification of traits with at least one significant association in the HLA region by shared pathophysiology.

While the role of HLA in infectious disease and autoimmunity is well-established, its role in other disease types is less clear. As such, we sought to quantify the enrichment of association signal stratified by disease groups. We classified the 1,035 diseases that had at least one GWAS association into 45 trait categories based on ICD codes. We then calculated the average per SNP enrichment of association signals for each disease category by comparing the number of independent associations inside the HLA region to the number in the rest of the genome (Supplementary Table 1).

Overall, we found a 17x enrichment in the HLA region relative to the rest of the genome averaged across all 1,035 diseases that had at least one GWAS association anywhere in the genome. The individual disease category with the highest enrichment was the Infectious trait group, with a 396x enrichment relative to the rest of the genome (Figure 2b). The overall enrichment across all diseases remained relatively unchanged (16.6x) even after excluding all infectious traits. In addition, the majority of other trait groups, including groups such as Dental traits (71x), Dermatologic traits (63x enriched), Rheumatic traits (53x enriched), Hematologic (50x enriched), and Ear traits (45x enriched) also showed a major enrichment in the HLA region. In contrast, the Congenital group was the only group not enriched in the HLA region. This could be because the traits in the Congenital group are oligogenic, with an average of 2.2 hits outside the HLA region and none within the HLA locus. The most enriched trait groups showed enrichment for primarily two reasons (Supplementary Figure S1). First, some traits had high enrichment because they had many associations across the genome, with proportionately even more associations in the HLA region, such as the Rheumatic traits. Alternatively a subset of the enriched traits did not have many associations overall, but the few associations they had were in the HLA region, such as the Infectious traits.

To ensure that our results were robust and not driven by the unusually high gene density or by differences in genotype array coverage of the HLA region, we repeated our analyses to identify per-gene and per-base pair enrichments. The results were qualitatively consistent, differing by factors of 0.48x and 2.4x respectively. Overall, these results emphasize the involvement of the HLA region in a broad range of disease groups, including those from a variety of different pathologic mechanisms and organ systems.

In order to understand how the HLA region contributes to disease mechanisms, we next examined traits that had associations within the locus (N = 572 diseases). To remove essentially redundant traits, we focused on the subset of these traits that had LDSC genetic correlation *≤* 0.95. This included 269 diseases and 3 continuous traits (height, weight, body mass index). We then used forward stepwise regression to identify conditionally independent SNP associations for each trait. This resulted in 428 associations (MAF > 1%, P < 10^−6^) across all traits.

Classifying disease categories by ICD code, as was done in the enrichment analysis above, primarily results in anatomical groups as opposed to groups based on shared pathophysiology. To understand the contribution of HLA to biological disease mechanisms, we manually classified the 269 HLA-associated diseases based on pathophysiology (Figure 2c, Supplementary Table 2). For traits where the underlying mechanism is unknown or ambiguous, we classified by the organ system affected.

We calculated the number of traits in each of these trait categories that had at least one significant HLA association in the HLA region (Figure 2c). Two of the top disease categories were Rheumatic (40 traits) and Infectious (38 traits). In contrast to the enrichment analysis, additional multi-system disease groups beyond Rheumatic and Infectious traits were well-represented, including Autoimmune (27 traits) and Cardiometabolic (27 traits).

### Pleiotropy and spatial structure of significant SNP association signal within the HLA region

We aimed to evaluate the spatial distribution of the significant SNP association signal across the HLA region. We first categorized the associations by assigning each variant to its nearest gene (Figure 3a). We observed association signals throughout the extended HLA region with the highest density of associations near the twelve classical HLA genes, particularly the class II genes. However, associations were spread broadly across the region, with a total of 75 genes that were the nearest gene for at least one association, 59 of which were non-HLA genes. Overall, the associations were spread relatively consistently across trait groups, although the autoimmune and rheumatic traits had slightly higher signal near the class I genes than the other trait groups did, likely driven at least in part by the well-known associations of *HLA-B* alleles with rheumatic traits [52, 53] (Supplementary Figure S2).

**Figure 3:**
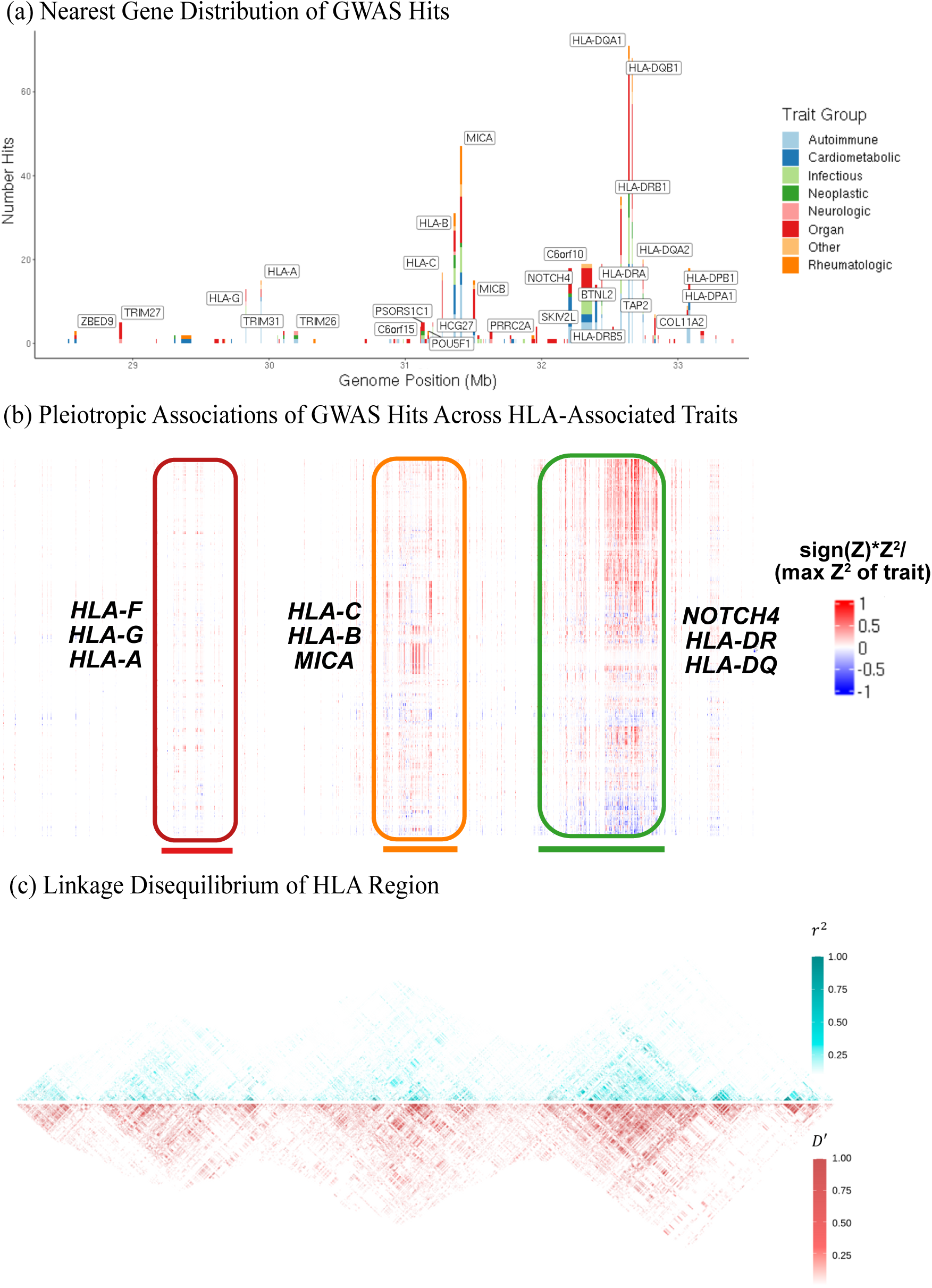
Pleiotropic structure of the HLA region. **A**. Distribution of significant SNP associations across the HLA region, binned by nearest gene. Each bar represents a different gene and the width corresponds to the length of the gene boundaries. **B**. Heatmap of normalized Z-scores for the 428 variants in the HLA region significantly associated with at least one trait. The x-axis corresponds to the genome position of the variant, the y-axis corresponds to the HLA-associated traits. Associations with all HLA-associated traits are shown for all variants that had an independent significant association with at least one trait. The three blocks used in subsequent analysis are circled, underlined, and labeled by well-known genes within each block. **C**. Linkage disequilibrium as measured by r and D^′^ of the approximately 40,000 SNPs covering the HLA region (MAF > 1%).

We next evaluated the role of genetic variation in the HLA region in modulating disease risk trade-offs. We calculated normalized Z-scores for each association discovered in the forward stepwise analysis (sign(Z)\**Z*^2^/(max *Z*^2^ of trait); See Methods), and visualized how these association signals were spread across the locus (Figure 3b). We found that 99% of the associations were also significant (P < 10^−6^) for one or more diseases beyond the trait for which they were identified as a conditionally independent significant association. Moreover, we found variants that significantly increased the risk for one disease while significantly decreasing risk for another disease suggesting a possible risk trade-off between traits.

The normalized Z-scores visually clustered around three main genomic regions within the HLA locus. The first cluster spanned two non-classical and one class I HLA gene (*HLA-F*, *HLA-G*, *HLA-A*). The second spanned two class I HLA genes and one non-HLA gene (*HLA-C*, *HLA-B*, *MICA*). The third spanned one non-HLA gene and two sets of class II HLA genes (*NOTCH4*, *HLA-DR*, *HLA-DQ*).

The overall pleiotropic structure revealed large blocks of SNPs spanning hundreds of kilobases that have similar effects across traits. These encompass multiple genes, and likely arise due to the high gene density and the extensive linkage disequilibrium (LD) in the region (Figure 3; Supplementary Figure S3).

### Pleiotropic disease associations at the haplotype level

The HLA region is particularly challenging for standard association studies because of its strong LD, multiallelic sites, and large effect coding variants within the classical HLA genes. Motivated by the block-like structure of the HLA locus (Figure 3b), we developed an approach to explore pleiotropy at the haplotype level, with haplotype blocks spanning multiple genes and including non-classical HLA, non-HLA, and non-coding regions.

The three main regions (“blocks”) described above were selected based on the density of signal from the significant SNP associations, overlapping LD patterns, and functional relevance. We defined haplotypes for each of the three regions by the unique combination of phased nucleotides at 1,000 randomly selected biallelic SNPs with MAF > 1% (Figure 4; see Methods for additional details). We then clustered related haplotypes into groups (Supplementary Table 3; Supplementary Information 1), and for each block performed association analyses between the haplotype groups and the 269 HLA-associated diseases. We discovered 469 significant trait-haplotype group associations (|Z| > 4) across blocks (Figure 5; Supplementary Table 4), representing 64 traits. Of these traits, 25 had significant associations with all three blocks. Celiac disease had the most trait-haplotype group associations with 36 total (8 in Block 1, 16 in Block 2, 12 in Block 3), followed by rheumatic disease prescriptions with 34, spondylopathies with 32, and iridocyclitis and type 1 diabetes with 25 each.

**Figure 4:**
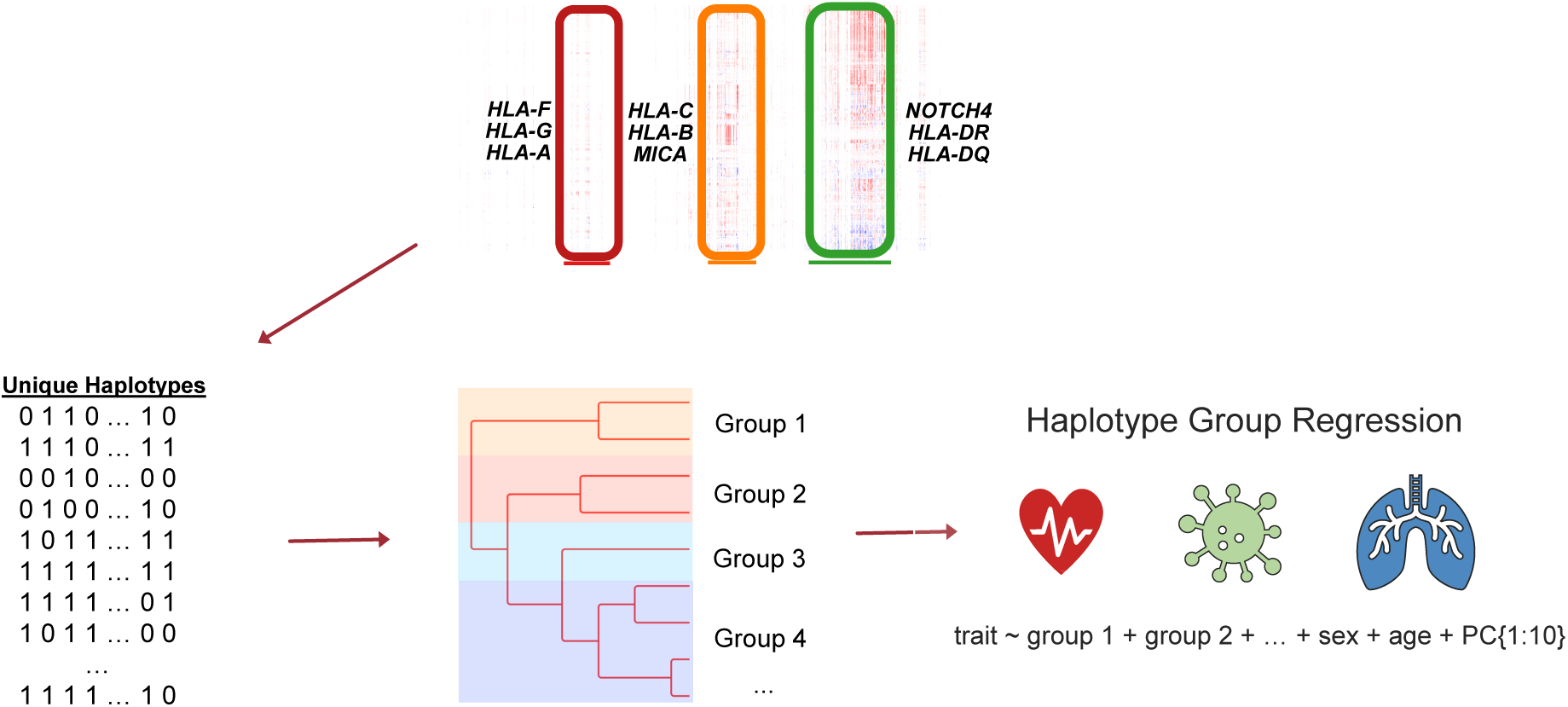
Haplotype group regression analysis pipeline. Overview of the pipeline for identifying the haplotype groups for each of the three blocks in the HLA region and performing trait associations. For each block, all unique phased combinations of nucleotides at 1,000 randomly selected SNPs were considered as haplotypes. We then clustered related haplotypes into groups by recursively splitting the dendrogram at each branch point (see Methods). Finally, for each of the three blocks, we performed association analyses between the haplotype groups and the 269 HLA-associated diseases, including all haplotype groups for a given block except the most frequent in each regression, as well as sex, age, and the first ten principal components of the genome-wide genotype matrix as covariates.

**Figure 5:**
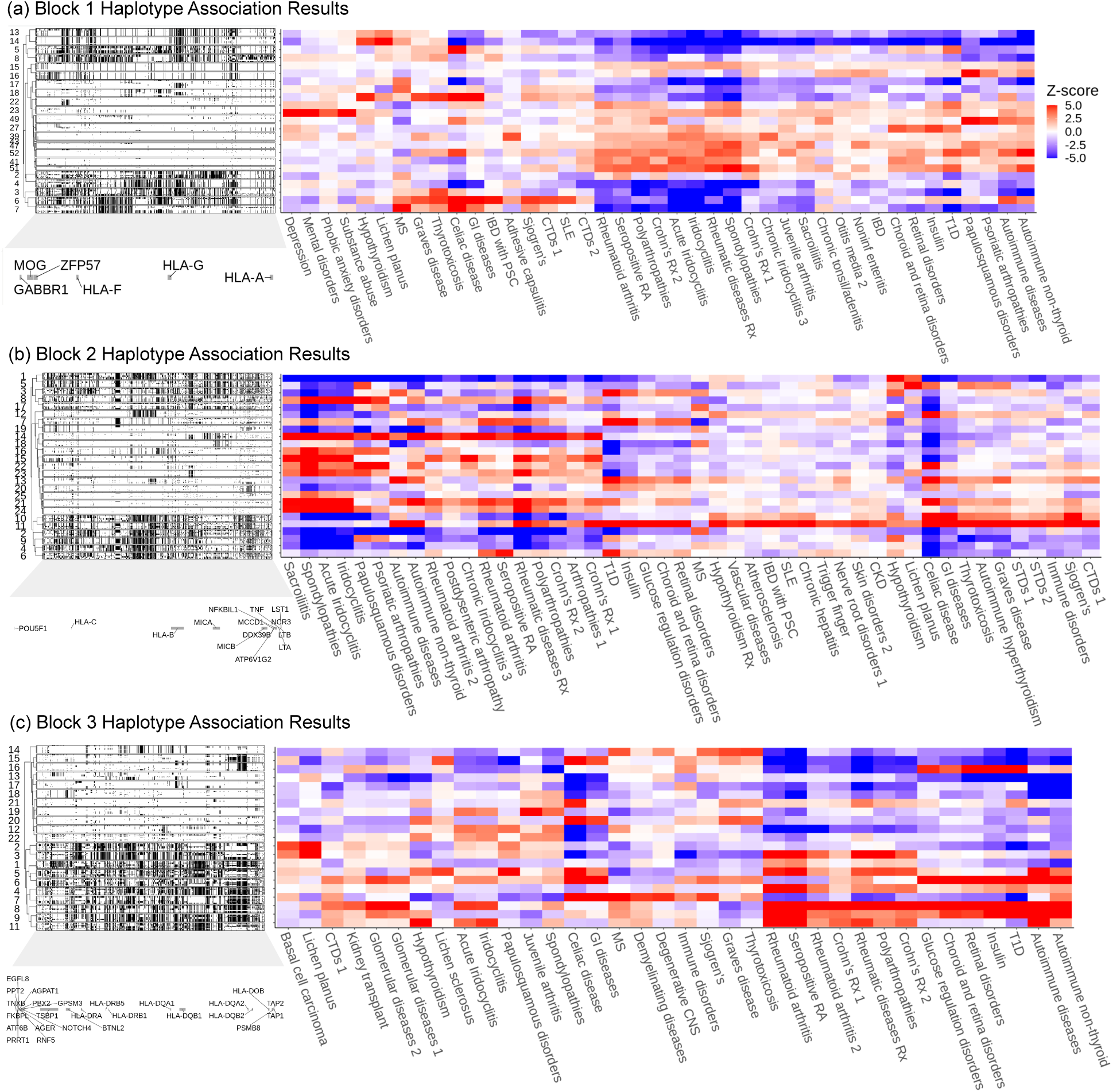
Haplotype group regression results. A dendrogram showing the clustering of the 40 most frequent haplotypes per haplotype group, with white representing the reference allele and black representing the effect allele. Genes are labeled below the corresponding SNPs overlapping their genome position, indicating which are within gene boundaries and which are intergenic. Heatmap showing the Z-scores from the haplotype group regression analysis across associated traits for **A**. Block 1, **B**. Block 2, and **C**. Block 3, including all traits with at least one association |Z| > 4 in that block, and all haplotype groups with at least one trait association or total copies greater than the minimum cutoff of 20,000 copies. For visualization purposes, traits are clustered and Z-scores were set to a maximum of |Z| of 5.

We sought to explore the patterns of pleiotropy within these blocks. For each block, we considered all traits with at least one association (|Z| > 4) in that block, and all haplotype groups with at least one trait association or total copies greater than the minimum cutoff of 20,000 copies (Figure 5). This resulted in 41 traits and 23 haplotype groups for Block 1, 46 traits and 25 haplotype groups for Block 2, and 36 traits and 21 haplotype groups for Block 3.

The majority of the haplotype groups were significantly associated with multiple traits. A subset of haplotype groups were associated with increased risk for some diseases, but decreased risk for others, consistent with disease risk trade-offs. For example, in Block 1, haplotype group 6 is associated with increased risk (Z > 3) for 10 traits, including GI autoimmune disorders, thyroid conditions, and connective tissue and rheumatic disorders. However, this haplotype group is also associated with decreased risk for 8 traits, mostly other rheumatic and inflammatory traits (Supplementary Table 4). Overall, of the 58 haplotype groups that showed a significant disease association (|Z| > 4), the mean number of associations (|Z| > 3) per haplotype group was 5 risk-increasing associations, and 7 risk decreasing associations (Supplementary Figure S4; Supplementary Information 2).

In contrast to these haplotype groups showing disease risk trade-offs, we also observed that some haplotype groups had the same direction of effect across the majority of associated traits (Figure 5). For example, haplotype group 49 in Block 1 was one of the rarest haplotype groups (0.09% frequency), but all 6 of the diseases with which it was significantly (|Z| > 3) associated were in the risk increasing direction, including depression and phobic anxiety disorders. This finding motivated us to calculate overall disease burden proportions for each haplotype group (Supplementary Figure S5). We defined the set of relevant diseases for each block as any disease that was significantly associated with at least one of the haplotype groups in that block. Then for each haplotype group in a given block, we identified the proportion of individuals in the haplotype group that had a diagnosis of at least one of the block’s relevant diseases. To identify the overall disease proportion as a baseline comparison, for each block we identified the proportion of all 412,181 individuals that had a diagnosis of at least one of the block’s relevant diseases. We then compared the haplotype group disease proportion to the overall disease proportion (Supplementary Figure S6). For example, compared to the baseline prevalence in FinnGen of 67.5%, we found that haplotype group 49 in Block 1 had one of the highest block-relevant disease burdens with 73% of carriers having at least one of the block’s significantly associated (|Z| > 4) diseases (P = 0.001). Our findings indicate that while some haplotypes had trade-offs in which diseases they increased and decreased the risk of, other haplotypes had an overall net positive or net negative impact across traits.

### Comparison of effects on trait pairs across haplotype groups

Many diseases have shared underlying pathology resulting in comorbidity. As a result, we expected to see sharing of associations across these diseases for the HLA haplotype groups. Indeed, our analysis recapitulated shared pathology for many traits, such as rheumatoid arthritis and seropositive rheumatoid arthritis, with similar associations across haplotype groups. More broadly we found that the inflammatory and rheumatic traits, such as spondylopathies, iridocyclitis, polyarthropathies, and rheumatoid arthritis clustered together throughout the three blocks (Figure 5). This could result from phenotypic correlations, caused, for example, by being co-morbid. An alternative explanation is that these traits have a shared biological mechanism modulated by genetic variation in the HLA region. Finally, it is possible that these correlations are an artifact of long-range LD extending beyond the haplotypes.

In contrast, we observed a surprising lack of concordance for a subset of seemingly similar traits, such as IBD and “IBD with primary sclerosing cholangitis” (IBD with PSC) (Figure 5). IBD with PSC is an idiopathic chronic liver disease complication developed by a subset of IBD patients, in which the bile ducts become inflamed and scarred, causing liver damage. IBD and IBD with PSC have a genome-wide genetic correlation of 0.45 and have similar effects across haplotypes in Block 3 (Pearson’s correlation of 0.57, SE = 0.12, P = 0.005), suggesting a shared etiology (Supplementary Figure S7). However, the haplotype groups have essentially uncorrelated effects on the two diseases in Block 1 (Pearson’s correlation of 0.10, SE = 0.13, P = 0.47). In fact, some haplotype groups in Block 1, such as group 6, are associated with increased risk for IBD with PSC, but not IBD (Supplementary Figure S7).

The difference in haplotype group effects on IBD and IBD with PSC is particularly interesting because it is difficult for clinicians to predict which IBD patients will develop liver damage and the mechanism leading to this damage is unknown [54]. Thus, understanding which parts of the genome are associated with increased risk for both a disease and its complications—as opposed to loci that differentially affect a disease and its complications—may help us better understand the factors that modulate the risk of certain disease complications. Understanding these differences may help explain why individuals with the same disease can present with a wide range of symptoms and outcomes.

To better disentangle whether these pleiotropic associations were due to LD, comorbidity, or shared biological pathways, we quantified the genome-wide LDSC genetic correlation, phenotypic correlation, and Pearson’s correlation across haplotype group trait associations for all pairwise combinations of haplotype group-associated traits for each block (Figure 6a). We discovered 1,520 pairs of traits with genome-wide genetic correlations greater than 0.3 where both traits are significantly associated (|Z| > 4) with at least one block. Of these trait pairs, 408 have a correlation across haplotype group effects > 0.3 for all three blocks, and surprisingly 256 had a discordant correlation of less than –0.3 in at least one block. We also observed discordant association signals for diseases with previously well-defined genetic associations and with clinical impact [52, 55, 56], such as Graves Disease and Rheumatoid Arthritis (Figure 6b).

**Figure 6:**
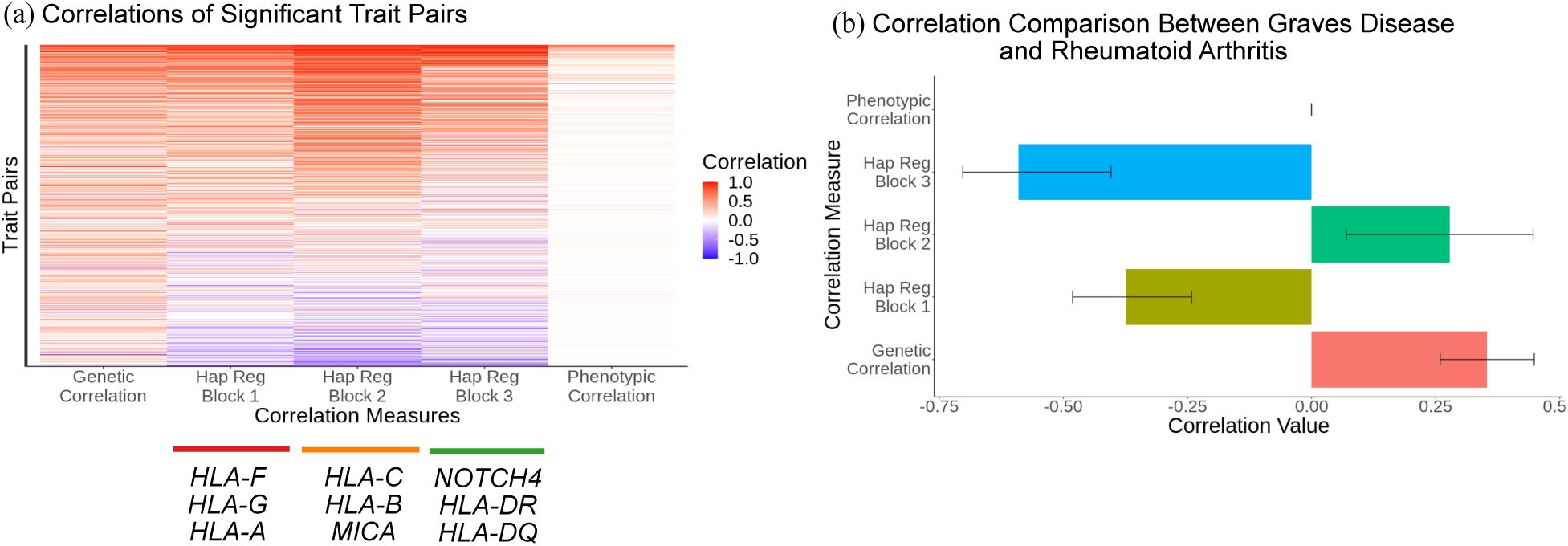
Correlation of haplotype associated traits. **A**. Overview and comparison of the pairwise relationships between traits that were significantly associated with the haplotype group regression analysis, comparing genome-wide LDSC genetic correlations, Pearson’s correlation across haplotype groups in each block, and phenotypic correlations. **B**. Comparison of correlation measures between Graves Disease and Rheumatoid Arthritis.

Graves disease is a condition where autoantibodies against the TSH receptor lead to overstimulation of the thyroid gland resulting in hyperthyroidism. Rheumatoid Arthritis is an idiopathic chronic inflammatory autoimmune disorder, primarily affecting the joints. Graves Disease and Rheumatoid Arthritis have a genome-wide genetic correlation of 0.35 (P = 0.0002), despite a phenotypic correlation of approximately 0 (P = 0.6). The correlation of effects within Block 2 is concordant—although not significantly so (Pearson’s correlation of 0.28, P = 0.18)—with this genome-wide genetic correlation. However, the effects in Blocks 1 and 3 are significantly negatively correlated (Pearson’s correlations of –0.37 and –0.59, P = 0.006 and 0.004 respectively). A potential explanation of this discordance between the genome-wide genetic correlation and the correlation within HLA regions is that these discordant regions affect a biochemical mechanism that breaks shared pathology, resulting in an increased risk in one trait while decreasing risk in another, when relevant variants elsewhere in the genome typically cause a shared increase or decrease risk in both traits. In previous work, we showed that such mechanisms can result in associations with opposite signs on the traits, in spite of a positive genome-wide genetic correlation, driven by variants acting at the shared biochemical pathways between both diseases [57].

### Evaluation of haplotype group signal independent of HLA alleles

While protein-coding variation within the HLA genes likely contributes significantly to the disease associations at the haplotype level, a feature of the haplotype analysis is that it includes genetic variation beyond coding variants in classical HLA genes, including non-classical HLA genes, non-HLA genes, and non-coding variation. Therefore, we sought to determine if the haplotype analysis was able to capture signal beyond the HLA alleles. To be conservative, we only considered signal entirely independent (directly, or indirectly due to LD) of the HLA alleles by performing the haplotype group regressions while including all classical HLA alleles (frequency > 1%) in each block as covariates. Overall, we found that 129 haplotype associations remained significant (|Z| > 4) after accounting for HLA allelic variation (Supplementary Figure S8; Supplementary Table 4), particularly for Block 1. Specifically, Block 1 had 171 significant associations across 48 unique traits in our original analysis, and 50 significant associations (|Z| > 4) across 18 unique traits after adjusting for the alleles.

This indicates that many associations cannot be explained by HLA allele variation or signal tagged by it, and demonstrates that the haplotype group analysis was able to pick up on disease associations that would have been missed in traditional allele association analysis. Block 1 overlapped only one classical HLA gene, *HLA-A*, suggesting that our haplotype regression approach may be particularly beneficial for regions of the HLA that cover non-classical HLA genes. Moreover, including the HLA alleles as covariates *increased* the strength of 42 significant haplotype-trait associations, indicating that the haplotypes explain some variation independent of that explained by the HLA alleles.

To further disentangle the information provided by haplotypes, SNPs, and HLA alleles, we performed association analyses at each of these levels separately. For the allele associations, we performed regressions using two approaches. The first approach used the standard method of including one allele per regression, while the second performed a multivariable regression of all alleles (variance inflation factor < 5) within a given block. The results of our association analyses at the haplotype, SNP, and HLA allele level on the full cohort across all 2,459 traits are available in Supplementary Tables 4-6. In total, we identified 7,649 associations and 647 HLA-associated traits across the combined association analyses. In particular, we identified 1,750 significant associations within the HLA locus in the haplotype analysis, including 27 traits not identified in the SNP or HLA allele analyses. These traits included non-organic psychotic disorders, otorrhagia, vascular dementia, and rectal cancers. This emphasizes that analyzing variation at the haplotype level provides orthogonal information about the role of the HLA region in disease.

## Discussion

In this work, we investigated how genetic variation throughout the HLA region associates with disease with a focus on broad pleiotropic patterns. We quantified the enrichment of association signal in the HLA region relative to the rest of the genome. We found a strong enrichment of disease associations across a broad range of disease groups and organ systems. Unsurprisingly, infectious traits were almost 400-fold enriched in the HLA region compared to the rest of the genome, in spite of infections making up a minority of the HLA-associated traits. We also found enrichment across multiple disease categories and organ systems including cardiovascular and neuropsychiatric diseases. Overall, these findings indicate HLA is a major locus for disease risk, not only for infectious diseases, but for diseases across many organ systems and etiologies.

Even with the extreme enrichment for infection-related associations, we expect that there is still substantially more information to be gleaned about the role of HLA in mediating infection. Our enrichment analysis controls for how well-powered a trait is by using the number of associations in the rest of the genome as a baseline. However, while we find a huge enrichment, the absolute number of total associations is small. Infectious traits are often under-reported in large biobank cohorts: identifying cases requires patients to seek care for the infection, followed by testing to confirm the specific pathogen. The infectious traits that we identified with the clearest signal tended to be those with more consistent reporting such as sexually transmitted infections. Therefore, our findings indicate that there is likely more signal for infectious traits that will be discovered with larger samples or more systematic reporting.

We performed disease association testing with SNPs, HLA alleles, and haplotypes to capture disease associations throughout the entire HLA region, including non-classical HLA genes and noncoding regions. We developed a haplotype analysis approach that includes genetic variation outside of the classical HLA alleles. While many diseases strongly associate with canonical HLA alleles, the HLA region harbours hundreds of genes, many of which also play an important role in immune response and other biological processes. Our haplotype approach discovered disease associations in the HLA region that remained after adjusting for classical HLA alleles, particularly in the region that overlaps more non-HLA and non-classical HLA genes.

Furthermore, we found some haplotype groups that displayed disease risk trade-offs, being protective for some diseases and risk-increasing for others. Meanwhile, we found some haplotype groups that were more consistently associated with increased disease burden across tested diseases. In addition, our haplotype analysis discovered that local genetic correlation, genome-wide genetic correlation and phenotypic correlation between trait pairs are not always concordant. This discordance suggests that the HLA region plays not only an important, but also a distinct role relative to the rest of the genome in contributing to the shared biology underlying these diseases.

In total we identified 7,649 significant trait associations across 647 unique diseases in the HLA region. Here, we highlight interesting patterns across these traits and example associations, but we have only begun to explore the thousands of disease associations generated by these analyses. Therefore we are releasing the association test results as a resource for future studies of the HLA region (Supplementary Tables 4-6). For example, our haplotype association results identify multiple traits or disease complications of previously unknown pathology that cluster with traits with known mechanism. It could be fruitful to use these clusters to generate hypotheses about the biology underlying idiopathic traits. In addition, the haplotypes present in FinnGen represent only a fraction of the genetic diversity present in the world. As more large cohort data continue to become available from regions around the world, future studies will benefit from application of these methods in other cohorts to study the HLA region as the haplotype level.

In conclusion, this work offers insights into the role of the HLA region in modulating the complex interplay between hundreds of diseases. Our findings highlight haplotype regression analysis as an additional approach for studying genetic variation in the region beyond the classical HLA alleles. Our results also provide insight into the nature of pleiotropy in the region and highlight novel pathological processes for not only infectious and autoimmune diseases typically associated with HLA, but also across a broad range of diseases.

## Methods

### Biobank samples and participants

The FinnGen study (see Supplementary Table 7 for full list of FinnGen contributors) is a large-scale genomics initiative that has analyzed over 500,000 Finnish biobank samples and correlated genetic variation with health data to understand disease mechanisms and predispositions. The project is a collaboration between research organisations, biobanks within Finland, and international industry partners. Here, we used data from FinnGen Data Freeze 10, which is comprised of samples from 412,181 Finnish individuals, 21,311,942 variants, and 2,459 traits.

### FinnGen Identification of SNP Associations

The summary statistics used in this study were generated using Regenie v2.2.4 and the FinnGen Regenie pipeline [58]. Current age or age at death, sex, genotyping chip, genetic relationship, and the first 10 principal components of the genome-wide genotype matrix were included as covariates [59]. Fine-mapping was performed using the SuSiE “Sum of Single Effects” model [60], excluding the HLA region. Further details are available at https://www.finngen.fi/en.

### Defining the HLA region

The HLA region was defined as 28,510,120-33,480,577 based on the Genome Reference Consortium Assembly Grch38.p14 (hg38) (https://www.ncbi.nlm.nih.gov/grc/human/regions/MHC). Protein coding genes were identified by overlapping FinnGen annotated genes with the protein coding gene file from HGNC (https://www.genenames.org/download/statistics-and-files). The LD plot represents linkage disequilibrium as measured by *r*^2^ and *D^′^* for 41,183 SNPs covering the HLA region. This set of SNPs corresponds to the subset of the 41,234 SNPs (MAF > 1%) within the HLA boundaries remaining after pruning with “plink –ld-window 999999 –ld-window-kb 1000 –ld-window-r2 0.1”.

### GWAS hit processing

GWAS results were filtered to include all traits with at least one hit in the HLA region with P < 10^−6^. LD score regression [61] was used to generate genetic correlation estimates, with relevant eur_*_ld_chr files downloaded from https://data.broadinstitute.org/alkesgroup/LDSCORE/. To remove essentially redundant traits, we further filtered to traits with LDSC genetic correlation < 0.95 with all remaining traits. We filtered to the most significant SNP (MAF > 1%) in the HLA region for each of the remaining traits. We then used stepwise forward conditional analysis with Plink2 (https://www.cog-genomics.org/plink/2.0/) for each trait to identify additional independent significant SNPs (MAF > 1%) in the HLA region with P < 10^−6^. A significance threshold of P < 10^−6^ was selected modified from the genome-wide significance threshold of 5*×*10^−8^ because here we are only considering SNPs in the HLA region.

In the conditional analysis, we considered only unrelated individuals, reducing the sample size to 259,802. We adjusted for age, sex and 10 principal components of the genome-wide genotype matrix. Z-scores were calculated from the GWAS results for the associations of the 428 hits in the HLA region with all the 272 HLA-associated traits. For visualizing effects across traits, we normalized squared Z-scores for each trait by the maximum *Z*^2^ for that trait. The sign of each SNP’s effects were assigned such that the SNP had a positive median Z-score across traits.

### Enrichment analysis

All traits with at least one associated SNP (MAF > 1% and P < 10^−6^) anywhere in the genome were included, and binned into trait groups. A threshold of P < 10^−6^ was chosen for ascertaining SNP associations in the genome outside HLA to conservatively match the significance threshold used to identify significant associations in the HLA region via the method described above. Enrichment was calculated for each trait group by dividing the number of independent hits per SNP in the HLA region by the number of independent hits per SNP outside the HLA region. For verification that this enrichment was not driven by SNP density, this process was also repeated using enrichment per genes and per base pair.

### Defining Haplotype Groups

Three regions (“blocks”) in the HLA region were selected based on the density of signal from the significant SNP associations, overlapping LD patterns, and functional relevance. The first block was defined as 100kb below the start of the gene boundary of *HLA-F* to 100kb past the end of the gene boundary of *HLA-A*, 29,622,820 to 30,045,616 (Grch38.p14) and contained 5,022 SNPs. The second block was defined as 100kb below the start of the gene boundary of *HLA-C* to 100kb past the end of the gene boundary of *MICB*, 31,168,798 to 31,611,071 (Grch38.p14) and contained 8,073 SNPs. The third block was defined as 100kb below the start of the gene boundary of *NOTCH4* to 100kb past the end of the gene boundary of *HLA-DQA2*, 32,094,910 to 32,847,125 (Grch38.p14) and contained 11,027 SNPs.

Each block was then subset down to 1,000 randomly selected biallelic SNPs with MAF > 1% due to computational constraints of the clustering process. Each individual’s two phased haplotypes at these 1,000 positions were identified. Haplotypes were clustered by first removing rare haplotypes (defined as < 10 total copies across all participants), generating a dendrogram, and recursively splitting the dendrogram at each branch point from the root toward the tips until the total number of haplotypes below each node was less than the maximum threshold (defined as 80,000 copies or the maximum in a single haplotype, whichever was greater). Once the haplotype groups were identified, the rare haplotypes were then added to the group with which they clustered.

### Performing haplotype regression analysis

Logistic regression was then performed separately for each block for each of the 269 diseases with at least one SNP association in the HLA region for all haplotype groups, leaving out the haplotype group with the highest frequency. Sex, age, and the first ten principal components of the genomewide genotype matrix were included as covariates. The left out haplotype group was then set to 0 and the Z-scores of the regression results were then rescaled for each trait to have a mean of 0. A significance threshold of |Z| > 4 was chosen based approximately on the Bonferroni correction for the number of regressions (one for each of the 269 diseases) for each block at a significance level of 0.05.

In a follow-up analysis, we additionally performed haplotype regression analysis for all traits regardless of whether there was a GWAS hit in the HLA region for that trait, and for these regressions we applied a more stringent significance threshold of P < 6.7 *×* 10^−6^ to account for the additional traits tested (2459 traits * 3 blocks).

### Analysis of haplotype regression results

Subsequent analyses investigating patterns of pleiotropy of these haplotype groups focused on only the subset of diseases with at least one association |Z| > 4 in that block and the subset of haplotype groups with at least one trait association or total copies greater than the minimum cutoff of 20,000 copies. For these analyses, a significant threshold of |Z| > 3 was chosen based on the Bonferroni correction for the number of regressions (41 traits for Block 1, 46 for Block 2, and 36 for Block 3) for each block at a significance level of 0.05.

To calculate the overall disease burden proportion for each haplotype group, we defined the set of relevant diseases for each block as any disease that was significantly associated with at least one of the haplotype groups in that block. Then for each haplotype group in a given block, we identified the proportion of individuals in the haplotype group that had a diagnosis of at least one of the block’s relevant diseases. An individual was considered to be in a haplotype group if they were a carrier for at least one haplotype in the haplotype group. To identify the overall disease proportion as a baseline comparison, for each block we identified the proportion of all 412,181 individuals that had a diagnosis of at least one of the block’s relevant diseases. We performed an exact binomial test to determine the significance of the disease burden for haplotype group 49 in Block 1 to the block’s baseline disease prevalence of 67.5%.

### Allele regression analysis

To determine the extent to which the haplotype group signal remained after adjusting for the classical HLA alleles, we reran the haplotype group regressions while adjusting for the HLA alleles in each block (frequency > 1% and variance inflation factor < 5). We performed Firth’s Bias-Reduced Logistic Regression for all haplotype groups and alleles for each block and each trait using logistf (https://cran.r-project.org/web/packages/logistf/index.html). We then compared the Z-scores from the regression before and after adjusting for the alleles, using |Z| > 4 for the significance threshold. A significance threshold of |Z| > 4 was chosen based approximately on the Bonferroni correction for the number of regressions (one for each of the 269 diseases) for each block at a significance level of 0.05.

We performed the allele associations on all traits, regardless of whether there was a GWAS hit in the HLA region, using two approaches with sex, age, and 10 PCs included as covariates. For the first approach, we performed logistic regression separately for each block and each trait with one allele included in each regression, with a significance threshold of P < 2 *×* 10^−7^. This threshold was chosen to account for the additional traits tested (2459 traits * 98 alleles). For the second approach, we modeled all alleles within a block together jointly after we iteratively removed one regression variable at a time until all remaining had variance inflation factor < 5 to minimize issues of multi-collinearity, and applied a significance threshold of P < 6.7 *×* 10^−6^. This threshold was chosen to account for the additional traits tested (2459 traits * 3 blocks).

### Ethics statement

Participants in FinnGen provided informed consent for biobank research based on the Finnish Biobank Act. Alternatively, separate research cohorts, collected before the Finnish Biobank Act came into effect (in September 2013) and the start of FinnGen (August 2017), were collected based on study-specific consents and later transferred to the Finnish biobanks after approval by Fimea (Finnish Medicines Agency), the National Supervisory Authority for Welfare and Health. Recruitment protocols followed the biobank protocols approved by Fimea. The Coordinating Ethics Committee of the Hospital District of Helsinki and Uusimaa (HUS) approved the FinnGen study protocol (number HUS/990/2017).

The FinnGen study is approved by the Finnish Institute for Health and Welfare (permit numbers: THL/2031/6.02.00/2017, THL/1101/5.05.00/2017, THL/341/6.02.00/2018, THL/2222/6.02.00/2018, THL/283/6.02.00/2019, THL/1721/5.05.00/2019 and THL/1524/5.05.00/2020), the Digital and population data service agency (permit numbers: VRK43431/2017-3, VRK/6909/2018-3, VRK/4415/2019-3), the Social Insurance Institution (permit numbers: KELA 58/522/2017, KELA 131/522/2018, KELA 70/522/2019, KELA 98/522/2019, KELA 134/522/2019, KELA 138/522/2019, KELA 2/522/2020, KELA 16/522/2020), Findata permit numbers (THL/2364/14.02/2020, THL/4055/14.06.00/2020, THL/3433/14.06.00/2020, THL/4432/14.06/2020, THL/5189/14.06/2020, THL/5894/14.06.00/2020, THL/6619/14.06.00/2020, THL/209/14.06.00/2021, THL/688/14.06.00/2021, THL/1284/14.06.00/2021, THL/1965/14.06.00/2021, THL/5546/14.02.00/2020, THL/2658/14.06.00/2021, THL/4235/14.06.00/2021), Statistics Finland (permit numbers: TK-53-1041-17 and TK/143/07.03.00/2020 (earlier TK-53-90-20) TK/1735/07.03.00/2021, TK/3112/07.03.00/2021) and the Finnish Registry for Kidney Diseases permission/extract from the meeting minutes on 4th July 2019.

The Biobank Access Decisions for FinnGen samples and data utilized in FinnGen Data Freeze 10 include: THL Biobank BB2017_55, BB2017_111, BB2018_19, BB_2018_34, BB_2018_67, BB2018_71, BB2019_7, BB2019_8, BB2019_26, BB2020_1, BB2021_65, Finnish Red Cross Blood Service Biobank 7.12.2017, Helsinki Biobank HUS/359/2017, HUS/248/2020, HUS/430/2021 §28, §29, HUS/150/2022 §12, §13, §14, §15, §16, §17, §18, §23, §58, §59, HUS/128/2023 §18, Auria Biobank AB17-5154 and amendment #1 (August 17 2020) and amendments BB_2021-0140, BB_2021-0156 (August 26 2021, Feb 2 2022), BB_2021-0169, BB_2021-0179, BB_2021-0161, AB20-5926 and amendment #1 (April 23 2020) and its modifications (Sep 22 2021), BB_2022-0262, BB_2022-0256, Biobank Borealis of Northern Finland (2017_1013, 2021_5010, 2021_5010 Amendment, 2021_5018, 2021_5018 Amendment, 2021_5015, 2021_5015 Amendment, 2021_5015 Amendment_2, 2021_5023, 2021_5023 Amendment, 2021_5023 Amendment_2, 2021_5017, 2021_5017 Amendment, 2022_6001, 2022_6001 Amendment, 2022_6006 Amendment, 2022_6006 Amendment, 2022_6006 Amendment_2, BB22-0067, 2022_0262, 2022_0262 Amendment), Biobank of Eastern Finland (1186/2018 and amendment 22§/2020, 53§/2021, 13§/2022, 14§/2022, 15§/2022, 27§/2022, 28§/2022, 29§/2022, 33§/2022, 35§/2022, 36§/2022, 37§/2022, 39§/2022, 7§/2023, 32§/2023, 33§/2023, 34§/2023, 35§/2023, 36§/2023, 37§/2023, 38§/2023, 39§/2023, 40§/2023, 41§/2023), Finnish Clinical Biobank Tampere MH0004 and amendments (21.02.2020 & 06.10.2020), BB2021-0140 8§/2021, 9§/2021, §9/2022, §10/2022, §12/2022, 13§/2022, §20/2022, §21/2022, §22/2022, §23/2022, 28§/2022, 29§/2022, 30§/2022, 31§/2022, 32§/2022, 38§/2022, 40§/2022, 42§/2022, 1§/2023, Central Finland Biobank 1-2017, BB_2021-0161, BB_2021-0169, BB_2021-0179, BB_2021-0170, BB_2022-0256, BB_2022-0262, BB22-0067, Decision allowing to continue data processing until 31st Aug 2024 for projects: BB_2021-0179, BB22-0067, BB_2022-0262, BB_2021-0170, BB_2021-0164, BB_2021-0161, and BB_2021-0169, and Terveystalo Biobank STB 2018001 and amendment 25th Aug 2020, Finnish Hematological Registry and Clinical Biobank decision 18th June 2021, Arctic biobank P0844: ARC_2021_1001.

## Data Availability

Data produced are available online at https://doi.org/10.5281/zenodo.12763469.

## Acknowledgements

We want to acknowledge the participants and investigators of the FinnGen study. We thank Alyssa Lyn Fortier, Mineto Ota, Roshni Patel, Matthew Aguirre, Tami Gjorgjieva, and other members of the Pritchard lab for helpful discussions. This work has been supported by the National Science Foundation Graduate Research Fellowship, Stanford’s Knight-Hennessy Scholars Program, and the Stanford Center for Computational, Evolutionary and Human Genomics (C.J.S), the Finnish Medical Foundation (S.S.), and Instrumentarium Science Foundation and Academy of Finland #340539 (H.M.O.). The FinnGen project is funded by two grants from Business Finland (HUS 4685/31/2016 and UH 4386/31/2016) and the following industry partners: AbbVie Inc., AstraZeneca UK Ltd, Biogen MA Inc., Bristol Myers Squibb (and Celgene Corporation & Celgene International II Sàrl), Genentech Inc., Merck Sharp & Dohme LCC, Pfizer Inc., GlaxoSmithKline Intellectual Property Development Ltd., Sanofi US Services Inc., Maze Therapeutics Inc., Janssen Biotech Inc, Novartis AG, and Boehringer Ingelheim International GmbH. The following biobanks are acknowledged for delivering biobank samples to FinnGen: Auria Biobank (www.auria.fi/biopankki), THL Biobank (www.thl.fi/biobank), Helsinki Biobank (www.helsinginbiopankki.fi), Biobank Borealis of Northern Finland (https://www.ppshp.fi/Tutkimus-ja-opetus/Biopankki/Pages/Biobank-Borealis-briefly-in-English.aspx), Finnish Clinical Biobank Tampere (www.tays.fi/en-US/Research_and_development/Finnish_Clinical_Biobank_Tampere), Central Finland Biobank (www.ksshp.fi/fi-FI/Potilaalle/Biopankki), Biobank of Eastern Finland (www.ita-suomenbiopankki.fi/en), Finnish Red Cross Blood Service Biobank (www.veripalvelu.fi/verenluovutus/biopankkitoiminta) and Terveystalo Biobank (www.terveystalo.com/fi/Yritystietoa/Terveystalo-Biopankki/Biopankki/). All Finnish Biobanks are members of BBMRI.fi infrastructure (www.bbmri.fi). Finnish Biobank Cooperative – FINBB (https://finbb.fi/) is the coordinator of BBMRI-ERIC operations in Finland. The Finnish biobank data can be accessed through the Fingenious® services (https://site.fingenious.fi/en/) managed by FINBB. This work was supported by NIH grants RO1HG008140 and R01AG066490 (to J.K.P.).

## Data and Code availability

Code for project data analysis, processing and visualization is available at https://github.com/courtrun/HLA_finngen. Data generated from this study are available at https://doi.org/10.5281/zenodo.12763469.

## Supplement

### Supplementary Figures

**Figure S1:**
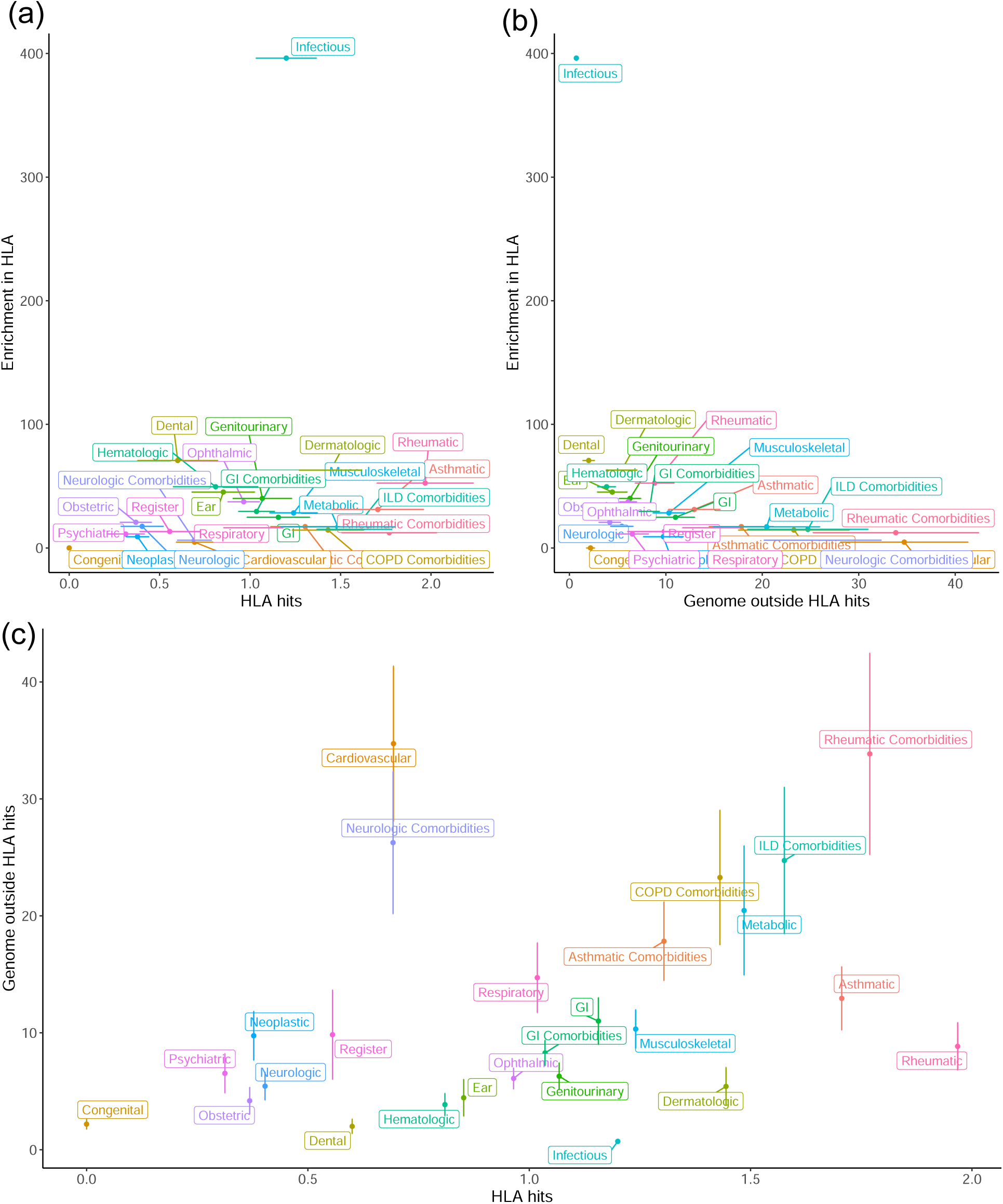
Enrichment by hits. Enrichment of fine-mapped GWAS hits in the HLA region relative to the number of hits throughout the genome outside the HLA region by trait group, compared to **A**. the number of HLA hits and **B**. the number of genome hits. **C**. Number of HLA hits versus the number of genome hits for each trait group.

**Figure S2:**
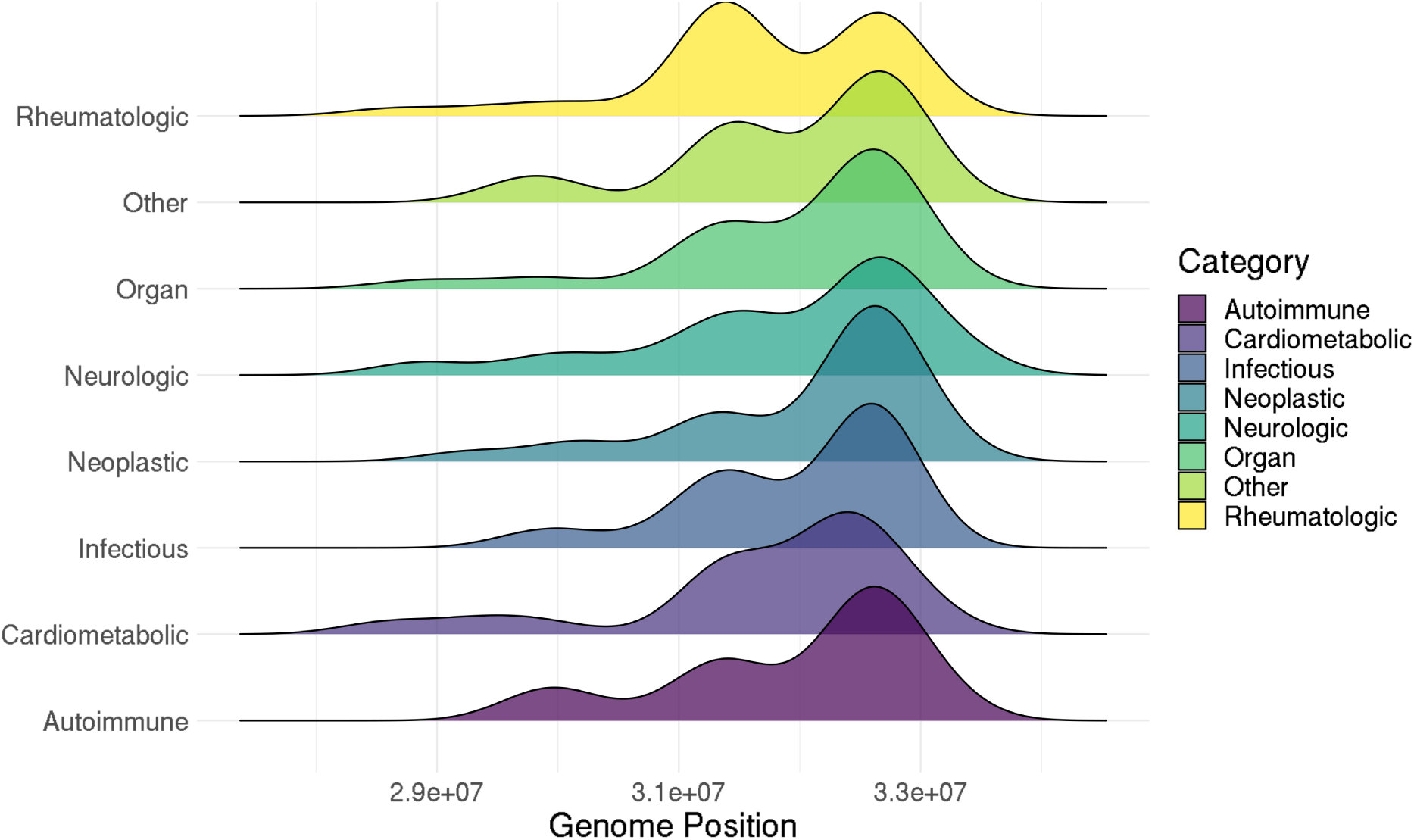
HLA distribution of significant SNP associations. Distribution of the significant SNP associations throughout the HLA region by trait group.

**Figure S3:**
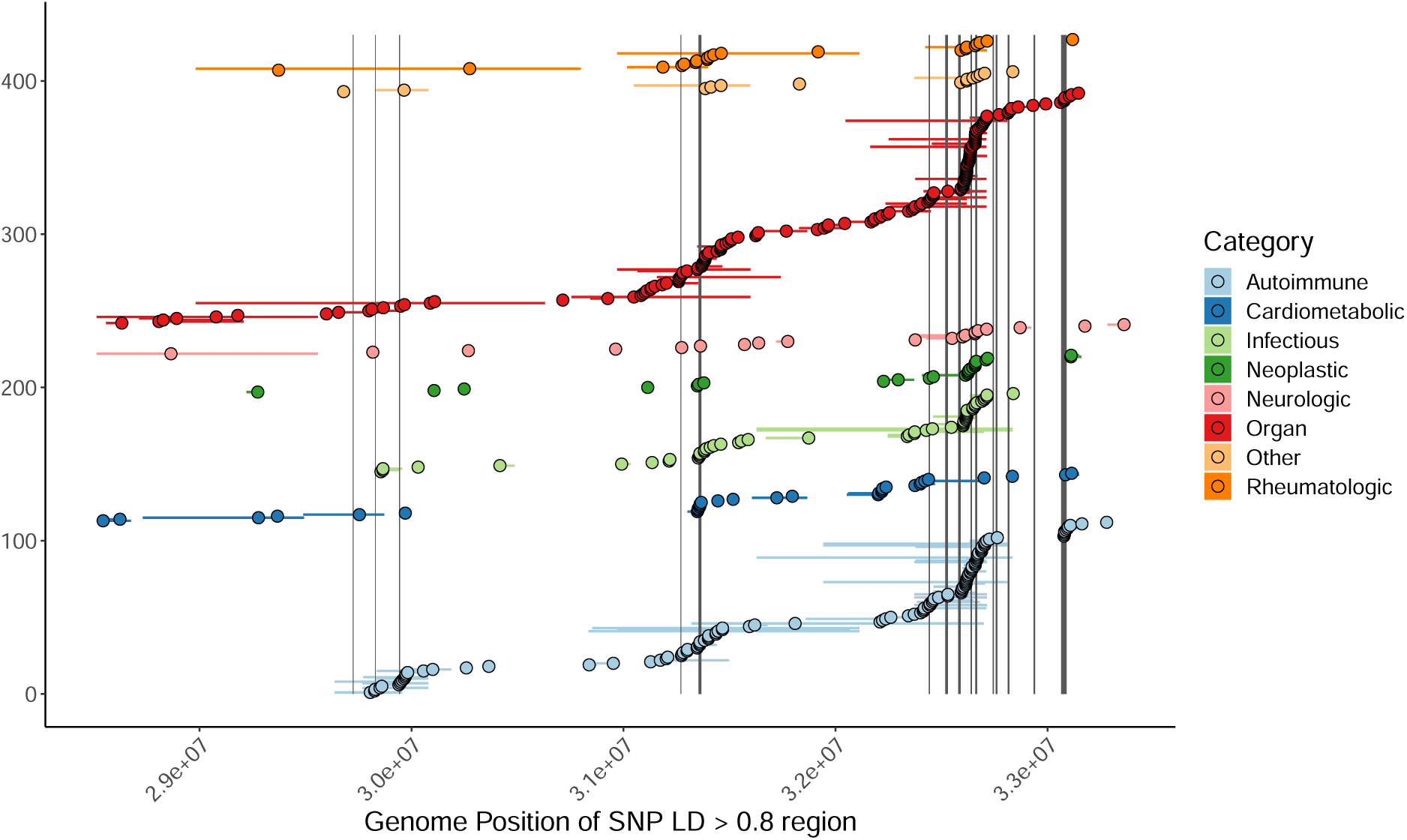
LD boundaries of hits. Genome position of the significant SNP associations in the HLA region with points corresponding to the SNP position and horizontal lines with the bounds corresponding to the lowest and highest genome position of SNPs in LD r^2^ > 0.8 with each hit.

**Figure S4:**
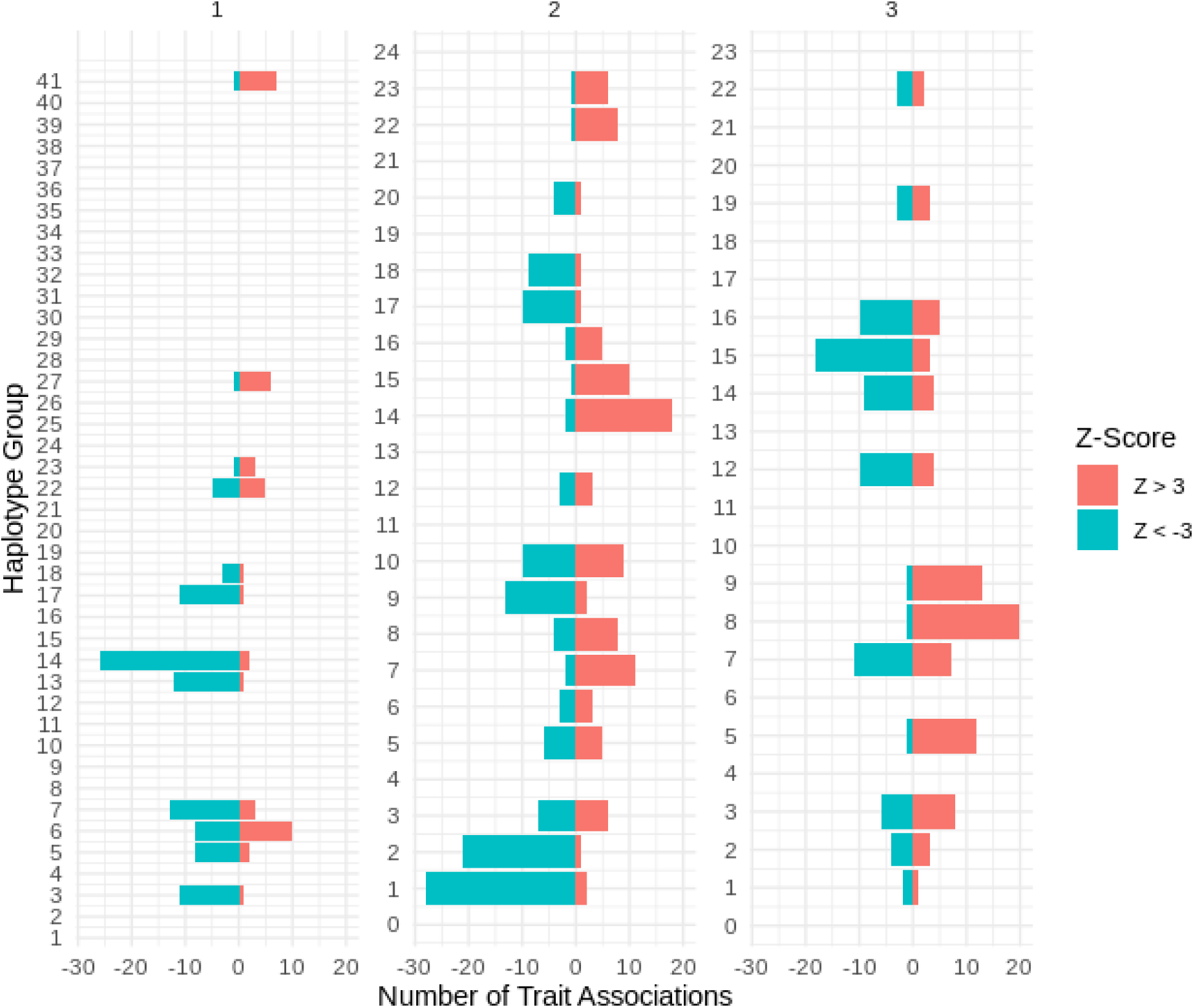
Haplotype group trait associations. Number of traits positively and negatively associated (|Z| > 3) with each haplotype group for all three blocks.

**Figure S5:**
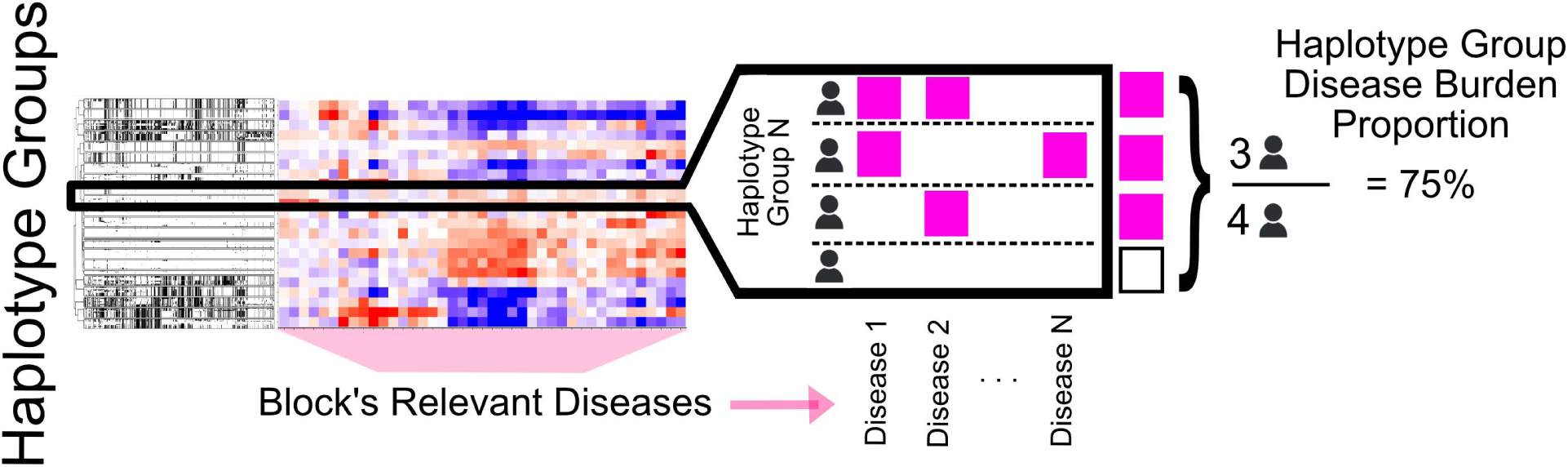
Disease burden analysis overview. Schematic of disease burden proportion analysis. For each haplotype group in a given block, the haplotype disease burden was defined as the proportion of individuals who were a carrier of at least one copy of a haplotype in the haplotype group that had a diagnosis of at least one of the block’s relevant diseases (example shown). For each block, the overall disease proportion across all individuals was calculated as the proportion of all individuals that had a diagnosis of at least one of the block’s relevant diseases.

**Figure S6:**
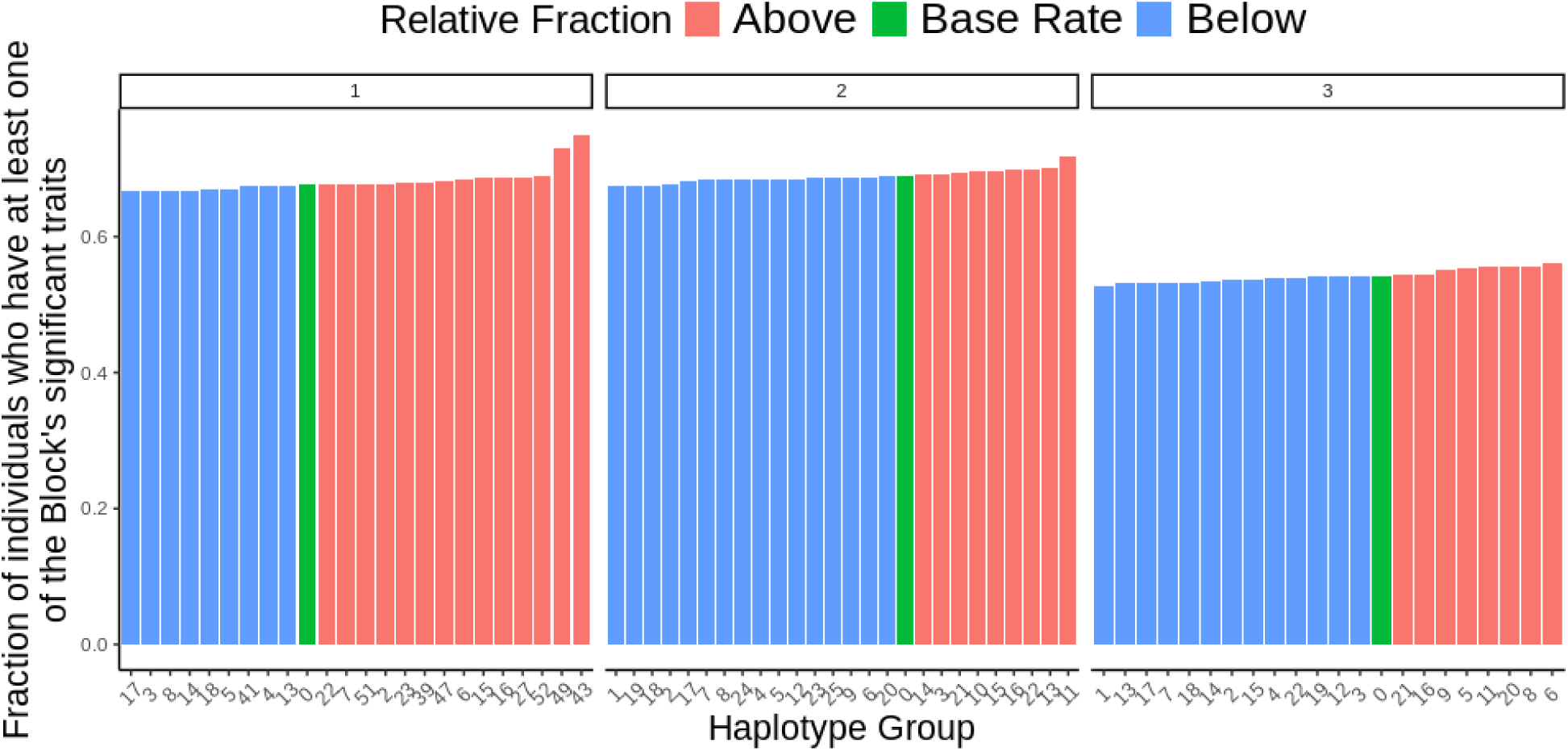
Haplotype group disease burden. Fraction of individuals in each haplotype group who had a diagnosis of at least one of the block’s significant traits, for each block. The overall disease proportion, or base rate, was defined as the fraction of individuals in all of FinnGen who had a diagnosis of at least one of the block’s significant traits and is shown in green. Haplotype groups with burden below the block’s base rate are in blue and those above are in red.

**Figure S7:**
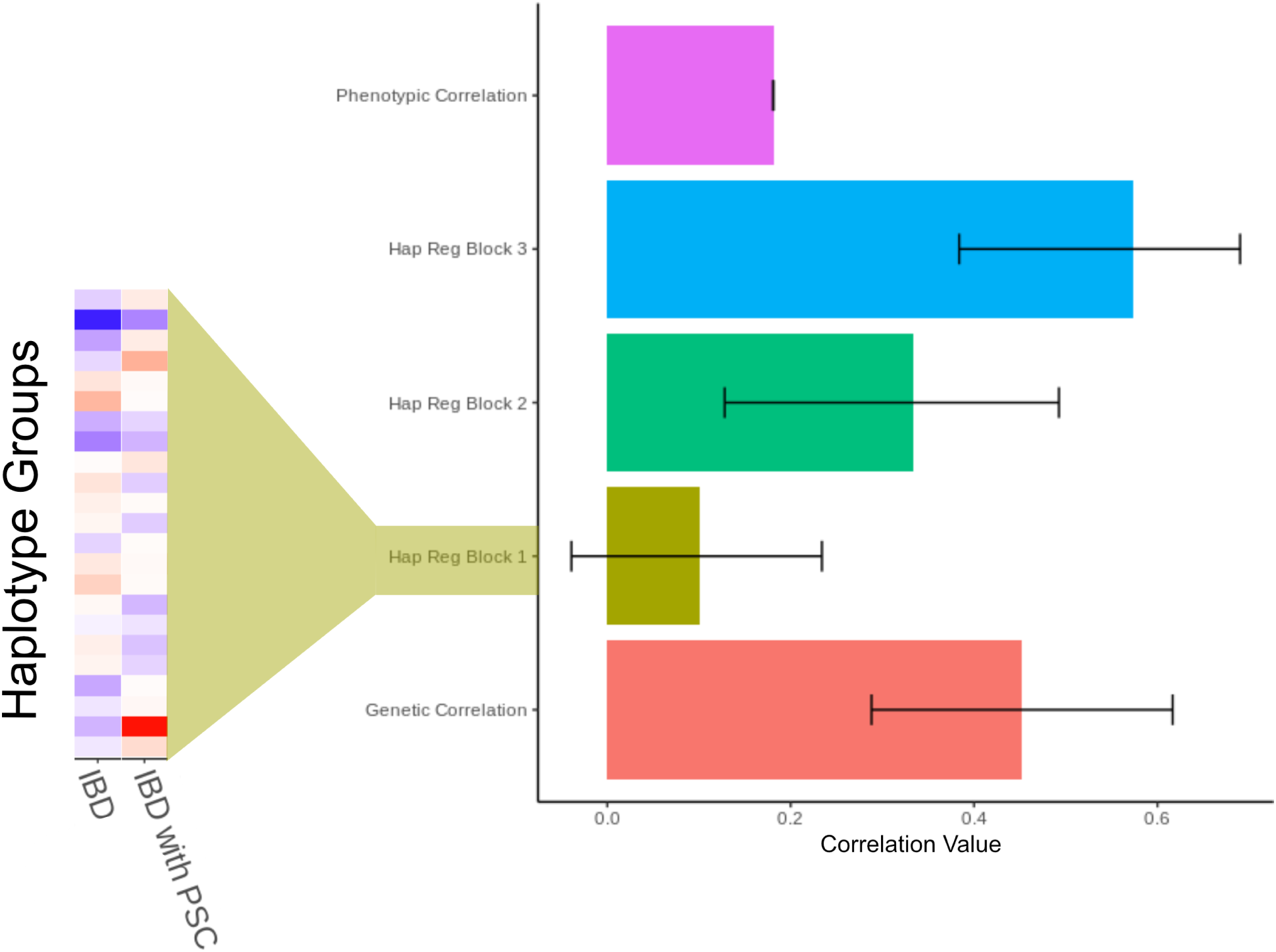
Comparison of IBD and IBD with PSC. Correlation measures between IBD and IBD with PSC. The inset for the haplotype group regression correlation for Block 1 corresponds to the Z-scores for individual haplotype groups in Block 1.

**Figure S8:**
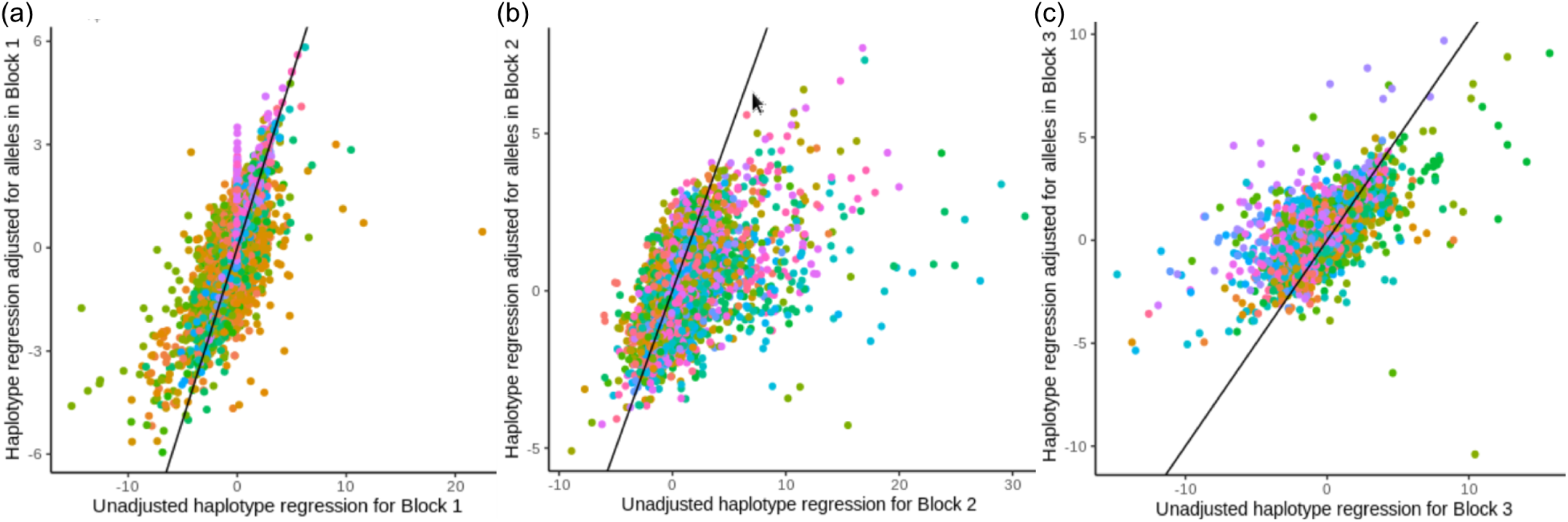
Allele adjusted haplotype group regressions. Comparison of the effects of the haplotype group regressions before and after adjusting for the classical HLA alleles across traits for each block. The points on the scatter plots correspond to Z-scores for different combinations of haplotype group trait associations.

### Supplementary Tables

Supplementary Table 1: Enrichment of GWAS hits in the HLA region for each trait group for all traits in FinnGen with at least one GWAS hit (MAF > 1%) anywhere in the genome.

Supplementary Table 2: Manual trait group classification for the 269 non-redundant diseases with at least one significantly associated SNP (P < 10^−6^; MAF > 1%) in the HLA region, by pathophysiology first (Category) and then by affected organ system (Subcategory).

Supplementary Table 3: Haplotype and haplotype group statistics and assignments. Haplotype statistics are for all haplotypes with > 10 total copies for privacy policy reasons.

Supplementary Table 4: Regression results for haplotype groups across all 3 blocks. The first tab has the data plotted in the heatmap of Figure 5, which is the values of the regression Z-scores rescaled to add back in the dropped haplotype group for each block. The next two tabs have the (non-rescaled) regression results for all traits, with and without jointly modeling with the relevant classical HLA alleles in the block.

Supplementary Table 5: Regression results for the SNP-trait associations for significant SNP associations remaining after step-wise conditional analysis in the HLA region.

Supplementary Table 6: Regression results for all allele associations for all traits, for both the approach jointly modeling alleles within a given block together (tab 1) and for the approach with one allele per regression (tab 2).

Supplementary Table 7: List of FinnGen contributors.

### Supplementary Information

Supplementary Information 1: Related haplotypes were clustered into haplotype groups for each block (Supplementary Table 3). This resulted in 53 haplotype groups for Block 1, with a mean of 15,554 total copies and a maximum of 81,997 copies (in haplotype group 15). Block 2 had 25 haplotype groups, with a mean of 32,974 copies and a maximum of 77,943 (in haplotype group 2). There were 22 haplotype groups for Block 3, with a mean of 37,471 copies and a maximum of 76,329 (for haplotype group 21). In Block 1, the mean number of trait associations per haplotype group was 6.25 traits, and haplotype group 14 had the maximum number of significant (|Z| > 4) associations with 25 trait associations. In Block 2, the mean trait associations per haplotype group was 8.4, and haplotype group 11 had the most with 30 trait associations. Block 3 had a mean of 6.8 trait associations per haplotype group, with haplotype group 8 having the maximum number of associations at 19. Across blocks, the mean number of significant trait associations for each of the block’s relevant haplotype groups was 7.2.

Supplementary Information 2: Multiple haplotype groups were positively associated with some traits and negatively associated with others. Haplotype group 22 in Block 1 is another example of a group with both positive and negative associations, including 8 traits with association Z > 2, and 13 with Z < –2. This haplotype group was associated with increased risk of Celiac disease, Graves disease and thyrotoxicosis. However, it was also associated with increased risk of hypothyroidism, Sjogren’s, and lichen planus. It was again negatively associated with traits like spondyopathies, iridocyclitis, rheumatoid arthritis, but also Type 1 diabetes, chronic tonsil/adenitis, and retinal disorders. In Block 2, haplotype group 10 is positively associated (Z>2) with 13 traits, including sexually transmitted diseases, chronic hepatitis, and Immune disorders. It is negatively associated with 14 traits, including many rheumatic disorders, as well as papulosquamous disorders, and psoriatic arthropathies. Similarly, haplotype group 7 of Block 3, is positively associated with 10 traits, such as multiple sclerosis, degenerative CNS disorders, and demyelinating diseases, and negatively associated with 16 traits, such as type 1 diabetes, retinal disorders, Lichen sclerosus, and juvenile arthritis.

## References

1. Horton, R., Wilming, L., Rand, V., Lovering, R. C., Bruford, E. A., Khodiyar, V. K., Lush, M. J., Povey, S., Talbot, C. C., Wright, M. W., et al. (2004). Gene map of the extended human MHC. Nature Reviews Genetics 5, 889–899. 10.1038/nrg1489.

2. Neefjes, J., Jongsma, M. L. M., Paul, P., and Bakke, O. (2011). Towards a systems understanding of MHC class I and MHC class II antigen presentation. Nature Reviews Immunology 11, 823–836. 10.1038/nri3084.

3. Ishigaki, K., Lagattuta, K. A., Luo, Y., James, E. A., Buckner, J. H., and Raychaudhuri, S. (2022). HLA autoimmune risk alleles restrict the hypervariable region of T cell receptors. Nature Genetics 54, 393–402. 10.1038/s41588-022-01032-z.

4. Fan, W.-L., Shiao, M.-S., Hui, R. C.-Y., Su, S.-C., Wang, C.-W., Chang, Y.-C., and Chung, W.-H. (2017). HLA Association with Drug-Induced Adverse Reactions. Journal of Immunology Research 2017. 10.1155/2017/3186328.

5. Parham, P. and Guethlein, L. A. (2018). Genetics of Natural Killer Cells in Human Health, Disease, and Survival. Annual Review of Immunology 36, 519–548. 10.1146/annurev-immunol-042617-053149.

6. Butler-Laporte, G., Farjoun, J., Nakanishi, T., Lu, T., Abner, E., Chen, Y., Hultström, M., Metspalu, A., Milani, L., Mägi, R., et al. (2023). HLA allele-calling using multi-ancestry whole-exome sequencing from the UK Biobank identifies 129 novel associations in 11 autoimmune diseases. Communications Biology 6, 1–17. 10.1038/s42003-023-05496-5.

7. Karnes, J. H., Bastarache, L., Shaffer, C. M., Gaudieri, S., Xu, Y., Glazer, A. M., Mosley, J. D., Zhao, S., Raychaudhuri, S., Mallal, S., et al. (2017). Phenome-wide scanning identifies multiple diseases and disease severity phenotypes associated with HLA variants. Science Translational Medicine 9. 10.1126/scitranslmed.aai8708.

8. Sakaue, S., Kanai, M., Tanigawa, Y., Karjalainen, J., Kurki, M., Koshiba, S., Narita, A., Konuma, T., Yamamoto, K., Akiyama, M., et al. (2021). A cross-population atlas of genetic associations for 220 human phenotypes. Nature Genetics 53, 1415–1424. 10.1038/s41588-021-00931-x.

9. Kennedy, A. E., Ozbek, U., and Dorak, M. T. (2017). What has GWAS done for HLA and disease associations? International Journal of Immunogenetics 44, 195–211. 10.1111/iji.12332.

10. Hurley, C. K. (2021). Naming HLA diversity: A review of HLA nomenclature. Human Immunology. Defining and Characterizing HLA Diversity 82, 457–465. 10.1016/j.humimm.2020.03.005.

11. Pierini, F. and Lenz, T. L. (2018). Divergent Allele Advantage at Human MHC Genes: Signatures of Past and Ongoing Selection. Molecular Biology and Evolution 35, 2145–2158. 10.1093/molbev/msy116.

12. Manczinger, M., Boross, G., Kemény, L., Müller, V., Lenz, T. L., Papp, B., and Pál, C. (2019). Pathogen diversity drives the evolution of generalist MHC-II alleles in human populations. PLOS Biology 17. 10.1371/journal.pbio.3000131.

13. Özer, O. and Lenz, T. L. (2021). Unique Pathogen Peptidomes Facilitate Pathogen-Specific Selection and Specialization of MHC Alleles. Molecular Biology and Evolution 38, 4376–4387. 10.1093/molbev/msab176.

14. Miyadera, H. and Tokunaga, K. (2015). Associations of human leukocyte antigens with autoimmune diseases: challenges in identifying the mechanism. Journal of Human Genetics 60, 697– 702. 10.1038/jhg.2015.100.

15. Radwan, J., Babik, W., Kaufman, J., Lenz, T. L., and Winternitz, J. (2020). Advances in the Evolutionary Understanding of MHC Polymorphism. Trends in Genetics 36, 298–311. 10.1016/j.tig.2020.01.008.

16. Takahata, N. and Nei, M. (1990). Allelic genealogy under overdominant and frequency-dependent selection and polymorphism of major histocompatibility complex loci. Genetics 124, 967–978. 10.1093/genetics/124.4.967.

17. Fortier, A. L. and Pritchard, J. K. (2022). Ancient Trans-Species Polymorphism at the Major Histocompatibility Complex in Primates. Preprint at bioRxiv.10.1101/2022.06.28.497781.

18. Arden, B. and Klein, J. (1982). Biochemical comparison of major histocompatibility complex molecules from different subspecies of Mus musculus: evidence for trans-specific evolution of alleles. Proceedings of the National Academy of Sciences 79, 2342–2346. 10.1073/pnas.79.7.2342.

19. Mayer, W. E., Jonker, M., Klein, D., Ivanyi, P., Seventer, G. van, and Klein, J. (1988). Nucleotide sequences of chimpanzee MHC class I alleles: evidence for trans-species mode of evolution. The EMBO Journal 7, 2765–2774. 10.1002/j.1460-2075.1988.tb03131.x.

20. Ohashi, A., Murayama, M. A., Miyabe, Y., Yudoh, K., and Miyabe, C. (2024). Streptococcal infection and autoimmune diseases. Frontiers in Immunology 15. 10.3389/fimmu.2024.1361123.

21. Hillary, R. P., Ollila, H. M., Lin, L., Desestret, V., Rogemond, V., Picard, G., Small, M., Arnulf, I., Dauvilliers, Y., Honnorat, J., et al. (2018). Complex HLA association in paraneoplastic cerebellar ataxia with anti-Yo antibodies. Journal of Neuroimmunology 315, 28–32. 10.1016/j.jneuroim.2017.12.012.

22. Santambrogio, L. and Marrack, P. (2023). The broad spectrum of pathogenic autoreactivity. Nature Reviews Immunology 23, 69–70. 10.1038/s41577-022-00812-2.

23. Bakkalci, D., Jia, Y., Winter, J. R., Lewis, J. E., Taylor, G. S., and Stagg, H. R. (2020). Risk factors for Epstein Barr virus-associated cancers: a systematic review, critical appraisal, and mapping of the epidemiological evidence. Journal of Global Health 10. 10.7189/jogh.10.010405.

24. Khan, G. and Hashim, M. J. (2014). Global burden of deaths from Epstein-Barr virus attributable malignancies 1990-2010. Infectious Agents and Cancer 9, 38. 10.1186/1750-9378-9-38.

25. Parkin, D. M. (2006). The global health burden of infection-associated cancers in the year 2002. International Journal of Cancer 118, 3030–3044. 10.1002/ijc.21731.

26. Bjornevik, K., Cortese, M., Healy, B. C., Kuhle, J., Mina, M. J., Leng, Y., Elledge, S. J., Niebuhr, D. W., Scher, A. I., Munger, K. L., et al. (2022). Longitudinal analysis reveals high prevalence of Epstein-Barr virus associated with multiple sclerosis. Science 375, 296–301. 10.1126/science.abj8222.

27. Bjornevik, K., Münz, C., Cohen, J. I., and Ascherio, A. (2023). Epstein-Barr virus as a leading cause of multiple sclerosis: mechanisms and implications. Nature Reviews Neurology 19, 160–171. 10.1038/s41582-023-00775-5.

28. Brown, M. A., Kenna, T., and Wordsworth, B. P. (2016). Genetics of ankylosing spondylitis– insights into pathogenesis. Nature Reviews Rheumatology 12, 81–91. 10.1038/nrrheum.2015.133.

29. Noble, J. A. and Valdes, A. M. (2011). Genetics of the HLA Region in the Prediction of Type 1 Diabetes. Current Diabetes Reports 11, 533–542. 10.1007/s11892-011-0223-x.

30. Ziade, N. (2023). Human leucocyte antigen-B27 testing in clinical practice: a global perspective. Current Opinion in Rheumatology 35, 235–242. 10.1097/BOR.0000000000000946.

31. Raiteri, A., Granito, A., Giamperoli, A., Catenaro, T., Negrini, G., and Tovoli, F. (2022). Current guidelines for the management of celiac disease: A systematic review with comparative analysis. World Journal of Gastroenterology 28, 154–175. 10.3748/wjg.v28.i1.154.

32. Amstutz, U., Shear, N. H., Rieder, M. J., Hwang, S., Fung, V., Nakamura, H., Connolly, M. B., Ito, S., Carleton, B. C., and CPNDS clinical recommendation group (2014). Recommendations for HLA-B*15:02 and HLA-A*31:01 genetic testing to reduce the risk of carbamazepine-induced hypersensitivity reactions. Epilepsia 55, 496–506. 10.1111/epi.12564.

33. D’Antonio, M., Reyna, J., Jakubosky, D., Donovan, M. K., Bonder, M.-J., Matsui, H., Stegle, O., Nariai, N., D’Antonio-Chronowska, A., and Frazer, K. A. (2019). Systematic genetic analysis of the MHC region reveals mechanistic underpinnings of HLA type associations with disease. eLife 8. 10.7554/eLife.48476.

34. Bettens, F., Ongen, H., Rey, G., Buhler, S., Sollet, Z. C., Dermitzakis, E., and Villard, J. (2022). Regulation of HLA class I expression by non-coding gene variations. PLOS Genetics 18. 10.1371/journal.pgen.1010212.

35. Dendrou, C. A., Petersen, J., Rossjohn, J., and Fugger, L. (2018). HLA variation and disease. Nature Reviews Immunology 18, 325–339. 10.1038/nri.2017.143.

36. Jin, Y., Roberts, G. H. L., Ferrara, T. M., Ben, S., Geel, N. van, Wolkerstorfer, A., Ezzedine, K., Siebert, J., Neff, C. P., Palmer, B. E., et al. (2019). Early-onset autoimmune vitiligo associated with an enhancer variant haplotype that upregulates class II HLA expression. Nature Communications 10, 391. 10.1038/s41467-019-08337-4.

37. Sekar, A., Bialas, A. R., De Rivera, H., Davis, A., Hammond, T. R., Kamitaki, N., Tooley, K., Presumey, J., Baum, M., Van Doren, V., et al. (2016). Schizophrenia risk from complex variation of complement component 4. Nature 530, 177–183. 10.1038/nature16549.

38. Gupta, A. and Thelma, B. K. (2016). Identification of critical variants within SLC44A4, an ulcerative colitis susceptibility gene identified in a GWAS in north Indians. Genes & Immunity 17, 105–109. 10.1038/gene.2015.53.

39. Zhang, X., Lucas, A. M., Veturi, Y., Drivas, T. G., Bone, W. P., Verma, A., Chung, W. K., Crosslin, D., Denny, J. C., Hebbring, S., et al. (2022). Large-scale genomic analyses reveal insights into pleiotropy across circulatory system diseases and nervous system disorders. Nature Communications 13, 3428. 10.1038/s41467-022-30678-w.

40. Ritari, J., Koskela, S., Hyvärinen, K., FinnGen, n., and Partanen, J. (2022). HLA-disease association and pleiotropy landscape in over 235,000 Finns. Human Immunology 83, 391–398. 10.1016/j.humimm.2022.02.003.

41. Bycroft, C., Freeman, C., Petkova, D., Band, G., Elliott, L. T., Sharp, K., Motyer, A., Vukcevic, D., Delaneau, O., O’Connell, J., et al. (2018). The UK Biobank resource with deep phenotyping and genomic data. Nature 562, 203–209. 10.1038/s41586-018-0579-z.

42. Hirata, J., Hosomichi, K., Sakaue, S., Kanai, M., Nakaoka, H., Ishigaki, K., Suzuki, K., Akiyama, M., Kishikawa, T., Ogawa, K., et al. (2019). Genetic and phenotypic landscape of the major histocompatibilty complex region in the Japanese population. Nature Genetics 51, 470–480. 10.1038/s41588-018-0336-0.

43. Mozzi, A., Pontremoli, C., and Sironi, M. (2018). Genetic susceptibility to infectious diseases: Current status and future perspectives from genome-wide approaches. Infection, Genetics and Evolution: Journal of Molecular Epidemiology and Evolutionary Genetics in Infectious Diseases 66, 286–307. 10.1016/j.meegid.2017.09.028.

44. Apps, R., Qi, Y., Carlson, J. M., Chen, H., Gao, X., Thomas, R., Yuki, Y., Del Prete, G. Q., Goulder, P., Brumme, Z. L., et al. (2013). Influence of HLA-C Expression Level on HIV Control. Science 340, 87–91. 10.1126/science.1232685.

45. Tian, C., Hromatka, B. S., Kiefer, A. K., Eriksson, N., Noble, S. M., Tung, J. Y., and Hinds, D. A. (2017). Genome-wide association and HLA region fine-mapping studies identify susceptibility loci for multiple common infections. Nature Communications 8, 599. 10.1038/s41467-017-00257-5.

46. Binder, M. D., Fox, A. D., Merlo, D., Johnson, L. J., Giuffrida, L., Calvert, S. E., Akkermann, R., Ma, G. Z. M., ANZgene, Perera, A. A., et al. (2016). Common and Low Frequency Variants in MERTK Are Independently Associated with Multiple Sclerosis Susceptibility with Discordant Association Dependent upon HLA-DRB1*15:01 Status. PLOS Genetics 12. 10.1371/journal.pgen.1005853.

47. Bosca-Watts, M. M., Minguez, M., Planelles, D., Navarro, S., Rodriguez, A., Santiago, J., Tosca, J., and Mora, F. (2018). HLA-DQ: Celiac disease vs inflammatory bowel disease. World Journal of Gastroenterology 24, 96–103. 10.3748/wjg.v24.i1.96.

48. Lundström, E., Gustafsson, J. T., Jönsen, A., Leonard, D., Zickert, A., Elvin, K., Sturfelt, G., Nordmark, G., Bengtsson, A. A., Sundin, U., et al. (2013). HLA-DRB1*04/*13 alleles are associated with vascular disease and antiphospholipid antibodies in systemic lupus erythematosus. Annals of the Rheumatic Diseases 72, 1018–1025. 10.1136/annrheumdis-2012-201760.

49. Canela-Xandri, O., Rawlik, K., and Tenesa, A. (2018). An atlas of genetic associations in UK Biobank. Nature Genetics 50, 1593–1599. 10.1038/s41588-018-0248-z.

50. Rioux, J. D., Goyette, P., Vyse, T. J., Hammarström, L., Fernando, M. M. A., Green, T., De Jager, P. L., Foisy, S., Wang, J., Bakker, P. I. W. de, et al. (2009). Mapping of multiple susceptibility variants within the MHC region for 7 immune-mediated diseases. Proceedings of the National Academy of Sciences 106, 18680–18685. 10.1073/pnas.0909307106.

51. Debebe, B. J., Boelen, L., Lee, J. C., IAVI Protocol C Investigators, Thio, C. L., Astemborski, J., Kirk, G., Khakoo, S. I., Donfield, S. M., Goedert, J. J., et al. (2020). Identifying the immune interactions underlying HLA class I disease associations. eLife 9. 10.7554/eLife.54558.

52. Busch, R., Kollnberger, S., and Mellins, E. D. (2019). HLA associations in inflammatory arthritis: emerging mechanisms and clinical implications. Nature Reviews Rheumatology 15, 364–381. 10.1038/s41584-019-0219-5.

53. Queiro, R., Morante, I., Cabezas, I., and Acasuso, B. (2016). HLA-B27 and psoriatic disease: a modern view of an old relationship. Rheumatology 55, 221–229. 10.1093/rheumatology/kev296.

54. Kim, Y. S., Hurley, E. H., Park, Y., and Ko, S. (2023). Primary sclerosing cholangitis (PSC) and inflammatory bowel disease (IBD): a condition exemplifying the crosstalk of the gut–liver axis. Experimental & Molecular Medicine 55, 1380–1387. 10.1038/s12276-023-01042-9.

55. Conigliaro, P., D’Antonio, A., Pinto, S., Chimenti, M. S., Triggianese, P., Rotondi, M., and Perricone, R. (2020). Autoimmune thyroid disorders and rheumatoid arthritis: A bidirectional interplay. Autoimmunity Reviews 19. 10.1016/j.autrev.2020.102529.

56. The China Consortium for the Genetics of Autoimmune Thyroid Disease (2011). A genome-wide association study identifies two new risk loci for Graves’ disease. Nature Genetics 43, 897– 901. 10.1038/ng.898.

57. Smith, C. J., Sinnott-Armstrong, N., Cichońska, A., Julkunen, H., Fauman, E. B., Würtz, P., and Pritchard, J. K. (2022). Integrative analysis of metabolite GWAS illuminates the molecular basis of pleiotropy and genetic correlation. eLife 11. 10.7554/eLife.79348.

58. Mbatchou, J., Barnard, L., Backman, J., Marcketta, A., Kosmicki, J. A., Ziyatdinov, A., Benner, C., O’Dushlaine, C., Barber, M., Boutkov, B., et al. (2021). Computationally efficient whole-genome regression for quantitative and binary traits. Nature Genetics 53, 1097–1103. 10.1038/s41588-021-00870-7.

59. Kurki, M. I., Karjalainen, J., Palta, P., Sipilä, T. P., Kristiansson, K., Donner, K. M., Reeve, M. P., Laivuori, H., Aavikko, M., Kaunisto, M. A., et al. (2023). FinnGen provides genetic insights from a well-phenotyped isolated population. Nature 613, 508–518. 10.1038/s41586-022-05473-8.

60. Wang, G., Sarkar, A., Carbonetto, P., and Stephens, M. (2020). A Simple New Approach to Variable Selection in Regression, with Application to Genetic Fine Mapping. Journal of the Royal Statistical Society 82, 1273–1300. 10.1111/rssb.12388.

61. Bulik-Sullivan, B. K., Loh, P.-R., Finucane, H., Ripke, S., Yang, J., Patterson, N., Daly, M. J., Price, A. L., and Neale, B. M. (2015). LD Score Regression Distinguishes Confounding from Polygenicity in Genome-Wide Association Studies. Nature Genetics 47, 291–295. 10.1038/ng.3211.

